# Distinct intrinsic neural connectivity of an emotion regulation network across the menopausal transition

**DOI:** 10.64898/2026.01.30.26345227

**Authors:** Franziska Weinmar, Ann-Christin S. Kimmig, Sofia Amaoui, Laura Gervais, Alkistis Skalkidou, Carmen Morawetz, Birgit Derntl

## Abstract

Menopause is a major psychoneuroendocrine transition which can impact emotional functioning and mental health. Although emotion regulation (ER) is fundamental for mental health, intrinsic neural connectivity supporting ER during the menopausal transition remains unexplored. Addressing this gap, this study provides the first examination of intrinsic effective connectivity within an ER-related network across menopausal stages. Resting-state fMRI data were acquired from 76 healthy premenopausal (n = 32), perimenopausal (n = 19), and postmenopausal (n = 25) women. Effective connectivity within a predefined ER network was examined using spectral dynamic causal modeling. Further, we assessed how intrinsic connectivity predicts self-reported ER ability within each group. While self-reported ER ability did not differ across groups, resting-state effective connectivity within the ER network varied in a stage-dependent manner, with most heterogeneous effects observed between pre- and perimenopause, suggesting a non-monotonic trajectory. Perimenopause was characterized by distinct changes in frontal interactions, reflecting a redistribution rather than a gradual shift toward postmenopausal connectivity. Differences regarding postmenopause were restricted to greater weighting of temporo-parietal network components. Connectivity–ER ability associations revealed stage-specific predictive profiles, with distributed fronto-temporal connectivity predicting ER ability in premenopause, frontal-restricted connectivity in perimenopause, and a single frontal connection with reversed predictive direction in postmenopause. Our findings demonstrate that comparable levels of trait-based ER ability are associated with divergent intrinsic network configurations rather than a uniform architecture. Identifying perimenopause as distinct transition window of intrinsic network organization advances hormone-sensitive models of intrinsic connectivity and provides a framework for understanding how baseline network organization may adapt during psychoneuroendocrine transitions in women.

## Introduction

Mental health is shaped by dynamic biopsychosocial influences [1,2], with psychological processes critically determining how biological and social factors are translated into cognitive and emotional responses [3]. Periods of major female hormonal transitions constitute sensitive windows in which these psychological processes are particularly challenged, increasing susceptibility to affective dysregulation [4,5]. One such transition is menopause, marked by profound changes in ovarian hormone signaling that interact with neural, cognitive, and psychosocial systems central to emotional functioning [6–9]. Accordingly, the menopausal transition is increasingly recognized as a critical period for mental health, characterized by elevated anxiety and depressive symptoms and increased risk for mood disorders (e.g., [9–12]). This highlights the influence of the psychoneuroendocrine transition to menopause on core emotional functions, including emotion regulation (ER), with implications for mental health [8,13–15].

Menopause is experienced by every female and marks the end of her reproductive capacity, while progressively reshaping systemic, neural and psychological functions [7,9]. Clinically, menopause is defined retrospectively, after 12 consecutive months without menstruation, and typically occurs between 49 and 52 years of age [16–18]. The spontaneous (vs. induced) transition to menopause, known as perimenopause, constitutes a dynamic phase of progressive loss of ovarian function, characterized by pronounced endocrine fluctuations and irregular menstrual cycles [17]. Its late stage is defined by at least two episodes of amenorrhea lasting over 60 days and extends into the first 12 months after the final menstrual period, when postmenopause begins [17]. Overall, perimenopause can last five to ten years on average and is characterized by rising follicle-stimulating hormone (FSH) and fluctuating, ultimately declining estradiol levels, reflecting reorganization of hypothalamic-pituitary-gonadal signaling [9,17,19,20]. These endocrine dynamics are paralleled by alterations in neural structure and function, including reduced glucose metabolism [21], gray and white matter volume loss (reviewed in [22–24]), increased amyloid-beta deposition [25,26] as well as changes in neural function (reviewed in [8,27,28]. Concurrently, perimenopausal neuroendocrine changes are accompanied by substantial health challenges. Up to 90% of women experience menopausal symptoms, including vasomotor and sleep disturbances, sexual dysfunction, urogenital atrophy, and broader cardiovascular, skeletal, and metabolic effects, often resulting in impaired quality of life [7–9,16,20,29,30].

In the face of increased physiological and psychosocial demands, ER is a fundamental psychological process influencing mental health [31]. ER is a multidimensional ability involving intrinsic and extrinsic processes to monitor, understand, and modulate emotional experiences [32]. ER is central for adaptive social interaction and general well-being; conversely, difficulties in ER are associated with the development and persistence of diverse psychopathologies, including mood disorders [31,33–35]. Previous functional neuroimaging studies have identified neural networks of ER that involve distributed regions across the prefrontal, temporal, and parietal cortices (for meta-analyses see [36–38]). These findings highlight interactions between cortical control regions and subcortical emotion-generating regions, consistent with models in which frontal and parietal areas modulate amygdala activity [39–41]. Within these networks, connectivity among dorsolateral, dorsomedial, ventrolateral, and ventromedial prefrontal cortices (PFC) and the amygdala is critical for adaptive regulation [42–44].

Building on this framework, emerging research shows that female hormonal transitions modulate both behavioral and neural ER. Across the menstrual cycle, fluctuations in ovarian hormones have been associated with adaptive regulation, with reduced efficiency reported during low-estradiol compared to high-estradiol phases [45–48]. In line with behavioral effects, estradiol modulates neural activity within regulation networks, with higher levels facilitating prefrontal recruitment during ER [49–51]. Complementing task-related findings, estradiol has been found to modulate effective connectivity within ER networks at rest, influencing both top-down regulatory and bottom-up emotion-generating circuits [52]. To demonstrate effects on effective connectivity, Derntl et al. [52] used spectral dynamic causal modelling (spDCM), a method that goes beyond resting-state functional connectivity by inferring directed causal influences between regions. By estimating hidden neural states from the cross-spectral density of blood-oxygenation-level-dependent (BOLD) signals, spDCM enables robust inference on large-scale directional coupling within and between intrinsic networks at rest [53,54]. Based on this method, enhanced inhibitory coupling in ER networks was observed at higher estradiol levels, interpreted as reduced neuronal effort and pointing to an intrinsic network state that facilitates adaptive ER [52]. Beyond ER-related circuits, menstrual cycle phase further modulates intrinsic connectivity dynamics of large-scale networks including the default mode, salience, and executive control systems [54–58]. Collectively, these findings highlight ovarian hormones as modulators of adaptive ER, including the connectivity of underlying neural networks during resting-state.

Despite accumulating evidence for dynamic interactions between hormonal transitions and ER, research during the menopausal transition remains scarce, particularly at the neural level. First task-based functional magnetic resonance imaging (fMRI) studies provide insights into neural correlates of ER in menopausal samples: Compared with younger cohorts, healthy peri- and postmenopausal women (N = 11) showed stronger dorsolateral PFC recruitment alongside reduced amygdala responses during ER tasks, suggesting increased reliance on top-down control during menopausal transition [59]. Similarly, pre-, peri-, and postmenopausal women (n = 15, 11, 28, respectively) showed greater recruitment of cognitive control regions during an ER task with advancing menopausal status, while perimenopausal women further exhibited a negative interpretation bias toward neutral stimuli [14]. These results suggest that menopausal stage is associated with changes in the neural implementation of ER, with perimenopause additionally linked to altered emotional interpretation. Converging evidence further links pronounced menopausal symptoms to greater ER difficulties, heightened affective burden, and reduced quality of life [60], whereas better ER during this period predicts lower depressive symptoms, greater resilience, and improved well-being [60,61]. Nonetheless, no study to date has examined resting-state effective connectivity within ER networks across the menopausal transition.

The present study addresses this gap by examining intrinsic ER network dynamics using spDCM [53], i.e., resting-state effective connectivity, in pre-, peri-, and postmenopausal women. Unlike task-based fMRI, resting-state effective connectivity captures intrinsic, task-independent network configurations that reflect preparatory control states and shape individual differences in ER capacity [62,63]. Specifically, we investigated (1) whether self-reported ER abilities differ across groups; (2) whether intrinsic neural dynamics differ within a predefined functional ER network [64] across menopausal groups; (3) how individual variability in self-reported ER ability modulates resting-state connectivity within each group; and (4) which network connections robustly predict self-reported ER ability within each group. Given the limited evidence on effective connectivity and its hormonal modulation [52,54], and despite associations between menopausal stages, altered emotional functions and mental health [13,14,59,60,61], evidence for changes in neural organization or trait-level ER in healthy women across the menopausal transition remains scarce. Accordingly, our hypotheses were exploratory. Nevertheless, we hypothesized that menopausal status may be associated with differences in resting-state effective connectivity and that intrinsic connectivity patterns would relate to self-reported ER ability.

## Methods

### Sample

Eighty-four women (35-59 years of age) participated in the study, of which eight participants were excluded form analyses (n = 4 due to incidental findings, n = 1 due to excessive movement/claustrophobia, n = 3 dropped out). Thus, the final sample included N = 76 women distributed across the following three groups:

**·** premenopausal women (PRE) with a natural menstrual cycle, measured during the early follicular phase, days 2-6 (n = 32, M_age_ = 42.19 ± 4.41 years)
**·** perimenopausal women (PERI) with menopausal symptoms and irregular menstrual cycles, i.e., at least two skipped cycles in the past two years but at least one cycle in the past 12 months, i.e., menopausal amenorrhea < 12 months (n = 19, M_age_ = 50.58 ± 4.17 years)
**·** postmenopausal women (POST) with menopausal amenorrhea ≥ 12 months (n = 25, M_age_ = 53.72 ± 3.02 years)

Prior menstrual cycle tracking confirmed a regular cycle duration of 25-35 days in the PRE, irregular cycles in PERI, and menopausal amenorrhea in the POST group. The hormone profiles of FSH, luteinizing hormone (LH), Anti-Müllerian hormone (AMH) are as expected for the respective hormonal phases and thus confirm the sample selection (see Table 1). Of the final sample, four PERI and two POST used combined menopausal hormone therapy (MHT) containing estradiol valerate and progesterone. The four PERI used transdermal estradiol in combination with oral progesterone, while the two POST used a combined oral estradiol and progesterone pill. One PERI participant additionally used transdermal testosterone as well as oral pregnenolone and dehydroepiandrosterone. Intake duration ranged from 2-80 months at the time of study inclusion. Participants eligible for the study were females of European descent between the ages of 35 and 60, who identified as women and were fluent in German. Exclusion criteria were magnetic resonance imaging (MRI) contraindication (e.g. metal implants), current or past mental, neurological or endocrine disorders (except for treated hypothyroidism with L-thyroxine), pregnancy or breast feeding in the last 12 months, a history of breast or ovarian cancer, hysterectomy or oophorectomy, and the use of hormonal contraceptives or copper intrauterine devices in the last six months. Participants were recruited through e-mail providers at the University of Tübingen, print- and social media advertisements as well as in social networks. All participants gave written informed consent, signed a data protection agreement prior to participation and received financial compensation. The study was approved by the Ethics Committee of the Medical Faculty of the University of Tübingen (413/2023BO2) and data were collected from 01/2024-10/2025.

**Table 1.**
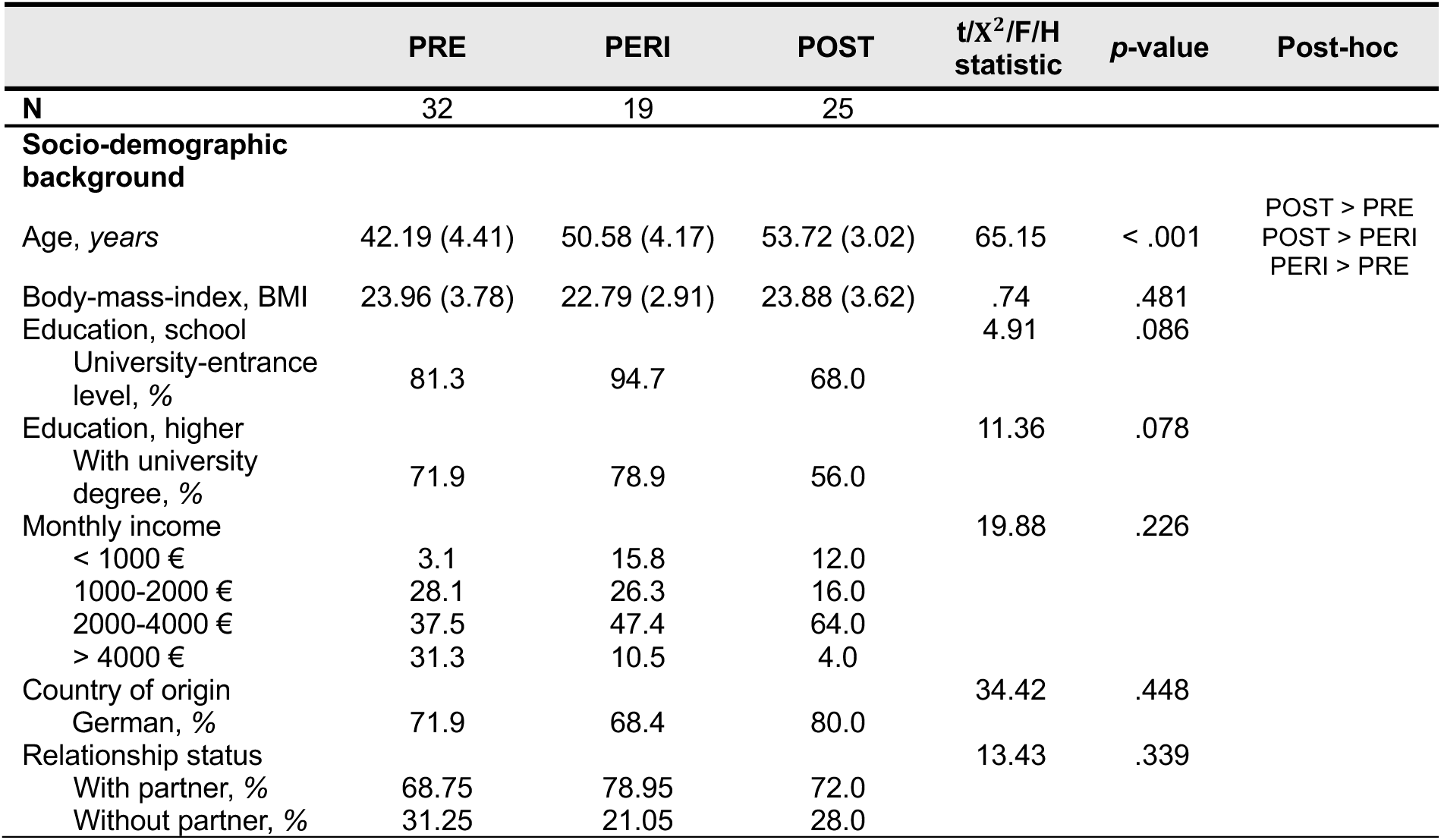
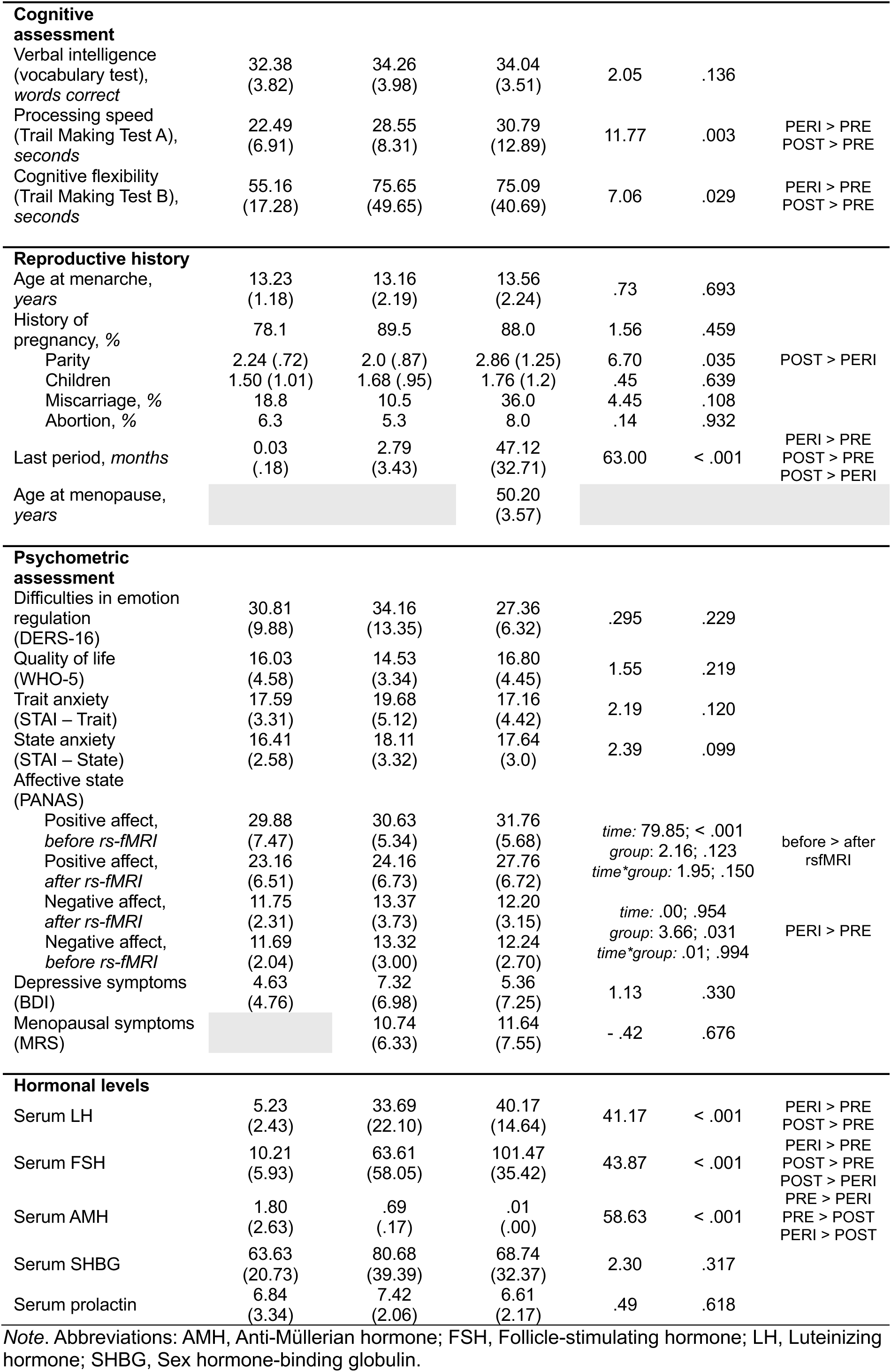
Sample description.

### Procedure

The study was part of a multimodal project on mental health focusing on reward processing, psychosexual health, and brain function in women across the menopausal transition. At an initial screening session, participants underwent a semi-structured clinical interview to assess exclusion criteria, including current or past mental disorders (Structured Clinical Interview for DSM-5 [65]) and MR-contraindications. Sociodemographic data and reproductive history were obtained, including menstrual cycle length (PRE), menopausal status (PERI, POST), previous use of hormonal contraceptives, pregnancies, and pregnancy losses. Use of MHT (type, regimen, dosage) was documented, if applicable. Cognitive screening included measures of verbal intelligence (Wortschatztest, WST [66]), processing speed, and cognitive flexibility (Trail Making Test, TMT [67]), to ensure cognitive functioning across participants. Participants also completed self-report questionnaires assessing depressive symptoms (Beck Depression Inventory-II, BDI-II [68]) as well as state and trait anxiety (State-Trait Anxiety Inventory, STAI [69]). Participants were subsequently invited to the fMRI session. Upon arrival, they completed self-report measures assessing current affect (Positive and Negative Affect Schedule, PANAS [70]) and state anxiety (STAI [69]), followed by additional questionnaires on menopausal symptoms (Menopause Rating Scale, MRS [71]) and emotion dysregulation (Difficulties in Emotion Regulation Scale-16, DERS-16 [72]; see below). MRI acquisition began with an anatomical scan, followed by resting-state fMRI (rs-fMRI), which was acquired as the first functional scan to avoid task-related carryover effects. Immediately after the rs-fMRI, affect ratings were repeated using the PANAS. Subsequent task-based fMRI and diffusion imaging are not reported here. At the end of the session, blood samples were collected by medical staff for hormone analyses (see below). PRE participants were invited for two fMRI sessions in counterbalanced order (early follicular phase: 2-6 days after menstruation onset; peri-ovulatory phase: ±2 days around ovulation), with cycle phase verified by menstrual tracking and ovulation testing. For the present study, only early follicular data from the PRE group were analyzed to minimize variability due to cyclical hormone fluctuations and to enhance comparability with the PERI and POST groups, which are characterized by lower endogenous hormone levels. Perimenopausal participants were scheduled, when feasible, during the early follicular phase based on estimated cycle timing; however, irregular cycles and ≤ 12 months of amenorrhea often precluded precise phase determination. POST participants were measured at the earliest convenient time point.

### Difficulties in Emotion Regulation Scale-16

Trait-based ER ability was measured with the Difficulties in Emotion Regulation Scale-16 (DERS-16 [72]), a 16-item short form of the Difficulties in Emotion Regulation Scale (DERS [73]). The DERS-16 assesses trait-level emotion dysregulation using five subscales: 1) Lack of emotional *clarity*, e.g., “I have difficulty making sense out of my feelings”; 2) *Non-acceptance* of emotional responses, e.g., “When I’m upset, I become angry at myself fore feeling that way”; 3) *Impulse* control difficulties, e.g., “When I’m upset, I become out of control”; 4) Difficulty in engaging in *goal-directed behavior*, e.g., “When I’m upset, I have difficulty getting work done”; 5) Limited access to ER *strategies*, e.g., “When I’m upset, I believe there is nothing I can do to feel better” [72,74]. Scores range from 16 to 80, with higher scores indicating greater emotion dysregulation [72,73]. The DERS-16 has shown strong psychometric properties, including high internal consistency, test-retest reliability, and convergent and discriminant validity [72].

### Hormone sampling and assessment

Hormone levels were assessed from blood samples obtained on the measurement day. Serum samples were analyzed at the central laboratory of the University Hospital Tübingen using enzyme-linked immunoassay (ELISA) to determine levels of LH, FSH, AMH, and prolactin from 7.5ml of blood. Assay sensitivities and measurement ranges were as follows: LH, 0.07-200.00mIU/ml; FSH, 0.30-200.00mIU/ml; AMH, 0.01-23ng/ml); SHBG, 1.60-180.00nmol/l; and prolactin, 0.30-200.00ng/ml.

### fMRI data acquisition

MRI data were acquired at a 3T Siemens MAGNETOM Prisma_XR scanner using a 64-channel head coil at the University Hospital Tübingen. First, a T1-weighted anatomical image was acquired (MPRAGE; 208 sagittal slices, TR = 2400ms, TE = 2.22ms, TI = 1000ms, voxel size = 0.8×0.8×0.8mm, flip angle = 8°, distance factor = 50%, GRAPPA acceleration factor = 2, FOV = 256mm). To account for magnetic field distortions in echo-planar images (EPIs), a T2*-weighted B_0_ inhomogeneity fieldmap was acquired prior to the remaining recordings (68 slices, TR = 745ms, TE = 5.19/7.65ms, voxel size = 2.3×2.3×2.0mm, FOV = 220mm, flip angle = 60°). Subsequently, functional images were acquired using a multiband T2*-weighted EPI sequence (68 interleaved slices, multiband factor = 4, TR = 1400ms, TE = 30ms, voxel size = 2×2×2mm, FOV = 220mm, flip angle = 65°). During the resting-state scan, 428 volumes were recorded (10min) while participants watched the standardized resting-state Inscapes video [75] and were instructed to remain awake, refrain from focusing on specific thoughts, and allow thoughts to pass naturally. B_0_ fieldmaps and functional images were acquired parallel to the AC-PC line using Siemens AutoAlign (T C-20.0°).

### Data analyses

All statistical analyses were conducted using IBM SPSS Statistics (version 27.0), unless stated otherwise. Group differences in sample characteristics and hormone levels were examined with univariate analyses of variance (ANOVAs), including group (PRE, PERI, POST) as the between-subjects factor. When ANOVAs yielded significant main effects, Bonferroni-corrected post-hoc comparisons were applied. In cases where the assumption of normality was violated, nonparametric Kruskal-Wallis H tests were performed. Significant omnibus effects in these tests were followed with pairwise Mann-Whitney U tests. To assess differences in self-reported affect (PANAS) between groups before and after rs-fMRI acquisition, a mixed between-within-subjects ANOVA was conducted with group (PRE, PERI, POST) as the between-subjects factor and time (pre fMRI, post rs-fMRI) as the within-subject factor, using affect ratings as the dependent variable. The significance threshold was set at α = .05 for all analyses. Effect sizes for ANOVAs are reported as partial eta-squared (ηp^2^), and effect sizes for Kruskal-Wallis tests as epsilon-squared (*ɛ*^2^), interpreted using conventional benchmarks (small ≈ .01, moderate ≈ .06, large ≈ .14 [76]). Missing data were not imputed; participants with incomplete data were excluded from the corresponding analyses.

#### Neuroimaging data analyses

##### Structural control analyses

To rule out potential confounding effects of brain morphology, we assessed group differences in global and region-of-interest (ROI) gray matter measures. No age-independent group differences were observed in total intracranial volume, global gray or white matter volume, or ROI-level gray matter volume, and these measures were therefore not included as covariates in subsequent spDCM analyses. Full details of the structural control analyses are provided in the supplement.

##### fMRI preprocessing

Rs-fMRI scans were preprocessed using *fMRIPrep* 24.1.1 [77–78] (RRID:SCR_016216), which is based on *Nipype* 1.8.6 [79,80] (RRID:SCR_002502). Preprocessing included motion and distortion correction, co-registration to the T1-weighted anatomy, normalization to Montreal Neurological Institute (MNI) space, high-pass filtering (128s), and removal of nuisance signals (motion parameters, global, CompCor), with motion outlier volumes censored. Full details of preprocessing with *fMRIPrep* are provided in the supplement. Spatial smoothing with a 6mm Gaussian kernel (full width at half maximum) was applied subsequently using SPM12. Participant head motion was low, with mean displacement remaining below .35mm across the sample and no individual exceeding 3.4mm.

##### ROI selection and time-series extraction

For each participant, a first-level General Linear Model (GLM) was estimated, including nuisance regressors for head motion (3 translations and 3 rotations) as well as white matter and cerebrospinal fluid signals, each including first derivatives. ROIs for the effective connectivity analysis were derived from a meta-analysis on ER [64]. This meta-analysis identified regions that were consistently engaged during both up- and downregulation of emotion through cognitive reappraisal. The ER network (see Figure 1) comprised frontal, temporal, and parietal regions, including left inferior frontal gyrus (LIFG, MNI: x = -46, y = 28, z = -8), right inferior frontal gyrus (RIFG, MNI: x = 50, y = 30, z = -8), right middle frontal gyrus (RMFG, MNI: x = 42, y = 24, z = 40), left medial frontal gyrus (LMeFG, MNI: x = -4, y = 12, z = 62), left middle temporal gyrus (LMTG, MNI: x = -60, y = -38, z = -2), left superior temporal gyrus (LSTG, MNI: x = -42, y = -56, z = 24), and right supramarginal gyrus (RSMG, MNI: x = 58, y = -54, z = 38). From the preprocessed fMRI data, we extracted BOLD time series for each ROI as the principal eigenvariate of all voxels within a 6mm radius sphere centered on the peak MNI coordinate of that ROI.

**Figure 1.**
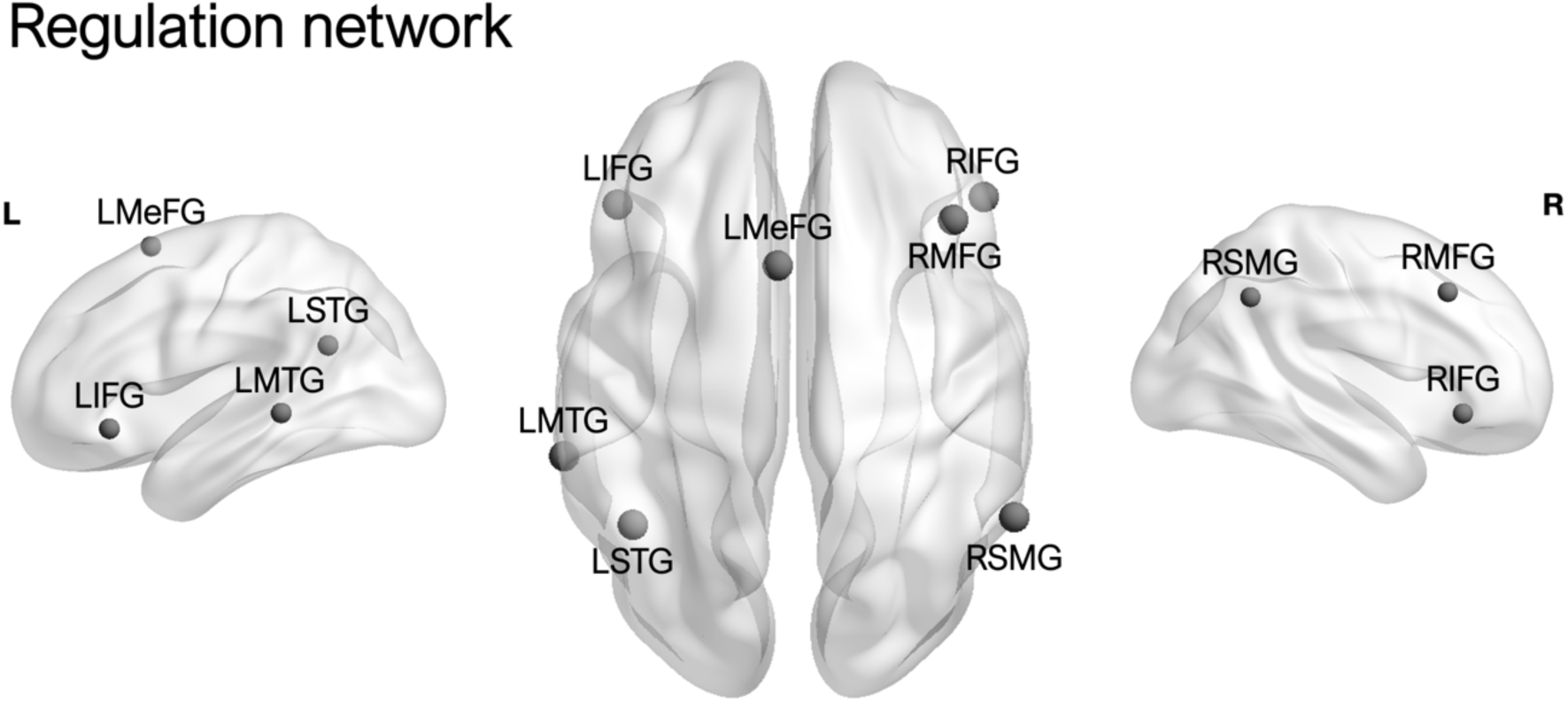
Regions of interest (ROIs) comprising the emotion regulation network. *Note*. Abbreviations: LIFG, left inferior frontal gyrus; RIFG, right inferior frontal gyrus; RMFG, right middle frontal gyrus; LMeFG, left medial frontal gyrus; LMTG, left middle temporal gyrus; LSTG, left superior temporal gyrus; RSMG, right supramarginal gyrus.

##### Spectral dynamic causal modeling

Effective connectivity among ROIs was estimated using spDCM implemented in DCM12 (SPM12). SpDCM models the cross-spectral density (second-order statistics) of endogenous neuronal fluctuations in the frequency domain, rather than the fMRI time series, thereby avoiding explicit estimation of latent neuronal states and enabling efficient deterministic model inversion under an assumption of stationarity [53]. At the first level, fully connected models of the regulation network were specified for each participant. These models were then inverted, meaning parameters were iteratively optimized to achieve the best fit to the observed data while respecting prior constraints [81]. Cross-spectral densities were modeled using a parameterized power-law representation of endogenous neuronal fluctuations [82]. The resulting estimates captured both directed interactions among regions and the amplitude of intrinsic neuronal activity within each ROI. Model inversion relied on a variational Laplace scheme [83]. Model evidence was quantified using variational Free Energy, which provides a lower bound on the (log) model evidence while guiding optimization of the posterior density. Model fit was high across participants, with the percentage of variance explained by the model for each participant ranging from 77.3% to 94.3%, indicating good data fit for all estimated models. Detailed diagnostics assessing model inversion quality for each participant are provided in Supplementary Figures S1 and S2.

##### Parametric empirical Bayes model

At the second level, the influence of menopausal status on effective connectivity within the regulation network was assessed using the hierarchical parametric empirical Bayes (PEB) framework for DCM [84]. Six separate PEB models were estimated. Three PEB models tested pairwise group differences in effective connectivity (PRE > POST, PRE > PERI, PERI > POST). Three additional PEB models examined associations between effective connectivity and individual differences in self-reported ER ability (DERS-16) within each menopausal group (PRE, PERI, POST). All PEB models included age as a covariate to account for age-related variance. Parallel PEB analyses excluding age as a covariate were conducted for the three pairwise group comparisons and are reported in the supplement, yielding broadly similar connectivity patterns. Bayesian Model Reduction (BMR) was applied to restrict the parameters and connectivity strengths to find the best model to explain the data. This exploratory approach assumes that all reduced models are equally probable a priori and discards those parameters that do not contribute to model evidence. The highest-ranking models were then combined through Bayesian model averaging (BMA), weighting parameter estimates by their evidence. Only effects with a posterior probability > .95 were considered robust and reported. To aid interpretability and avoid overemphasis of very small effects, only connections exceeding an absolute connectivity strength of ≥ |.10|Hz (and ≥ |.01|Hz for DERS-16 associations) were reported and visualized. All effects surpassing the posterior probability threshold, irrespective of their absolute connectivity strength, are fully reported in the supplement.

##### Prediction – cross-validation

In a final step, LOOCV was conducted to assess the predictive validity of effective connectivity parameters. For each group separately, a PEB model was fitted to all but one participant, and DERS-16 score was predicted for the left-out participant. This procedure was repeated over each participant and predictive performance was quantified as a Pearson correlation between the predicted an observed DERS-16 scores. Statistical significance was assessed with a *p*-value < .05.

#### Power considerations

For group comparisons in sample characteristics, sensitivity analyses (G*Power [85]) indicated 80% power to detect medium-to-large effects (f ≥ .36, ηp^2^ ≥ .115). In the present healthy sample, the observed group effect for DERS-16 was small (*ɛ*^2^ = .013), Accordingly, the study was not designed to detect subtle group differences in self-reported ER, but to examine whether intrinsic connectivity patterns vary across menopausal status and relate to individual differences in trait-based ER ability. For the spDCM analyses, classical power estimation is not applicable, as Bayesian frameworks rely on posterior probabilities and model evidence rather than null-hypothesis significance testing. Instead, the sample size was guided by evidence that DCM parameters are reliably estimated with n ≈ 20 per group [86]. Our smallest group (PERI, n = 19) meets this benchmark, and the PRE and POST groups exceed it. Robustness was supported by stringent posterior probability thresholds (> .95) and LOOCV-based predictive testing.

## Results

### Sample description

The three groups (PRE, PERI, POST) were comparable across most sociodemographic and psychometric variables. Significant group differences were observed for age (H = 97.17, *p* < .001, *ɛ*^2^ ≈ 1.0) and parity (H = 6.70, *p* = .035, *ɛ*^2^ = .077), with PRE being younger than PERI and POST (both *p*s < .001), PERI younger than POST (*p* < .001) and POST reporting more pregnancies than PERI (*p* = .024). As expected, time since last menstrual period differed across groups (H = 64.16, *p* < .001, *ɛ*^2^ = .852), increasing from PRE to PERI to POST (all *p*s < .001). Processing speed (H = 23.70, *p* < .001, *ɛ*^2^ = .297) and cognitive flexibility (H = 14.22, *p* < .001, *ɛ*^2^ = .167) were better in PRE than in PERI (both *p*s < .001) and POST (TMT-A: *p* < .001, TMT-B: *p* = .004) with latter groups showing no difference (TMT-A: *p* = .774, TMT-B: *p* = .553). However, all scores remained within age-adjusted normative ranges for the TMT [87], consistent with expectations for a cognitively healthy sample.

Positive affect decreased after rs-fMRI (*F*(1,73) = 79.85, *p* < .001, ηp^2^ = .522), with no group difference (*F*(2,73) = 2.16, *p* = .123, ηp^2^ = .056). Negative affect differed between groups (*F*(2,73) = 3.66, *p* = .031, ηp^2^ = .091), with higher scores in PERI than PRE (*p* = .026), while other groups did not differ (PRE vs POST: *p* = 1.0; PERI vs POST: *p* = .240). Negative affect did not change over time (*F*(1,73) = .00, *p* = .954, ηp^2^ = .00). No significant time-by-group interactions were observed for either positive or negative affect (both *ps* ≥ .150). A comprehensive overview of sample characteristics, psychometric measures, and hormone levels is provided in Table 1.

#### Hormone data

Groups differed in gonadotropic and ovarian hormones, confirming menopausal status: FSH levels (H = 88.34, *p* < .001, *ɛ*^2^ ≈ 1.0) were highest in POST, followed by PERI, and lowest in PRE (all *p* < .001). LH levels (H = 41.17, *p* < .001, *ɛ*^2^ = .55) were lower in PRE than in PERI and POST (both *p* < .001), with no difference between PERI and POST (*p* = .441). AMH levels (H = 58.63, *p* < .001, *ɛ*^2^ = .80) decreased from PRE to PERI to POST (all *p* < .001). No significant group differences emerged for SHBG or prolactin (*p* ≥ .153).

#### Emotion regulation

No significant group differences were observed in self-reported ER ability as measured by the DERS-16 (H = 2.95, *p* = .229, *ɛ*^2^ = .013). Overall scores reflected low levels of ER difficulties across groups (see Table 1), supporting its use as an index of trait-based ER ability in this sample.

### Neuroimaging results

#### Group differences in intrinsic regulation network connectivity

Second-level PEB analyses revealed strong evidence for group differences in intrinsic ER network effective connectivity across the menopausal transition (Figure 2). A concise overview of connectivity differences for each pairwise contrast is provided in Table 2 while thresholded connection estimates are reported in Table 3. The complete set of connections exceeding a posterior probability > .95 is provided in Supplementary Table S1 and visualized in Supplementary Figure S3. Positive values (green) indicate greater connectivity strength for Group A compared to Group B, whereas negative values (orange/red) denote reduced connectivity strength. Supplementary analyses additionally report results from models without age as a covariate.

**Figure 2.**
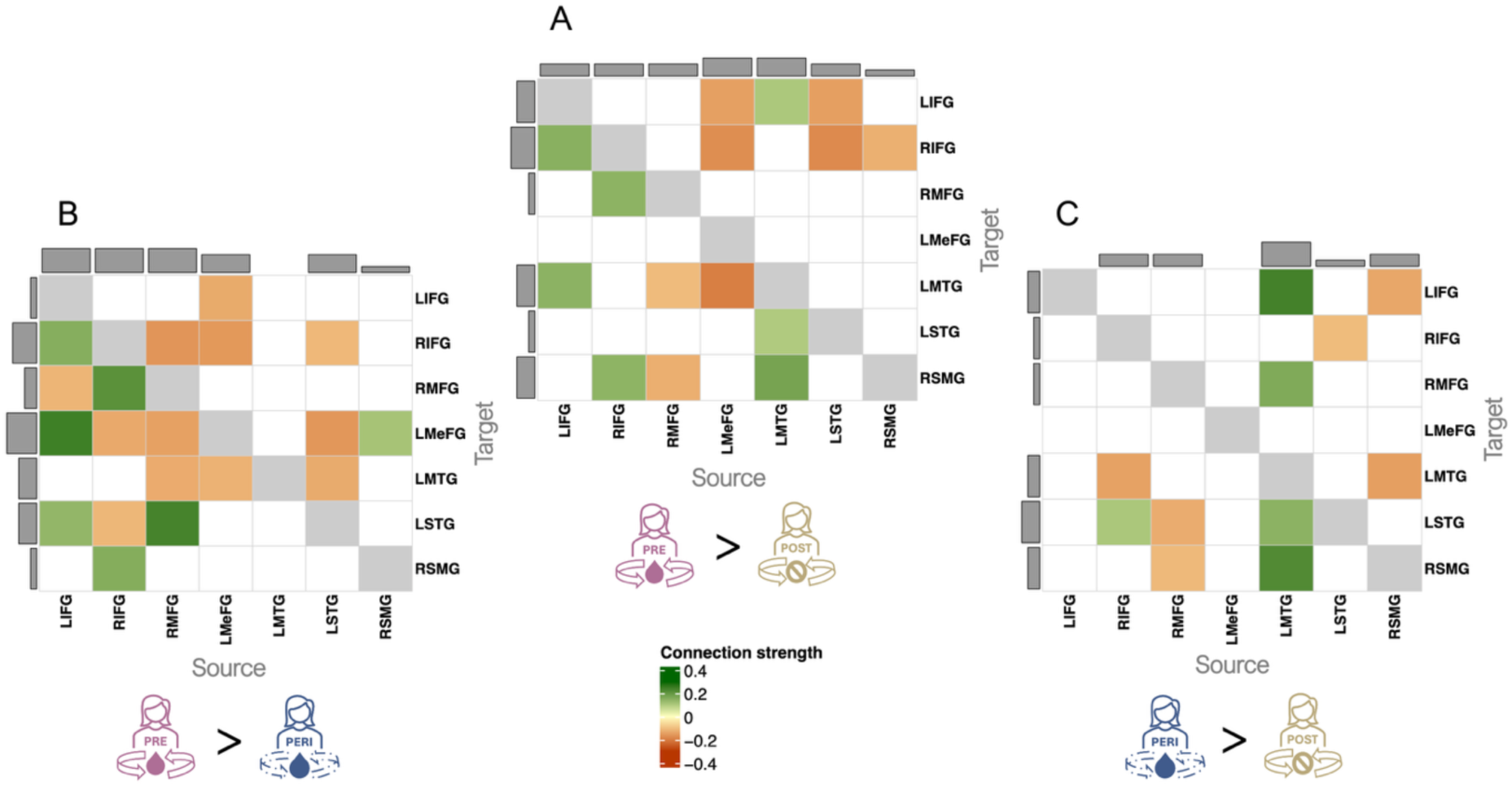
Connections showing strong evidence (posterior probability > .95) for group differences in effective connectivity across menopausal transition. (A) PRE > POST, (B) PRE > PERI, and (C) PERI > POST. Only connections with an absolute connectivity strength ≥ |.10| Hz are shown. *Note*. Displayed connections met both the posterior-probability criterion (PP > .95) and the absolute connectivity-strength threshold (|.10| Hz). Positive values (green) indicate greater connectivity strength for Group A compared to Group B, whereas negative values (orange/red) denote reduced connectivity strength. Effects reflect pairwise comparisons from PEB models including age as a covariate (PRE > POST, PRE > PERI, PERI > POST). Abbreviations: LIFG, left inferior frontal gyrus; RIFG, right inferior frontal gyrus; RMFG, right middle frontal gyrus; LMeFG, left medial frontal gyrus; LMTG, left middle temporal gyrus; LSTG, left superior temporal gyrus; RSMG, right supramarginal gyrus.

**Table 2.** Summary of group differences and self-reported emotion regulation ability-related associations in intrinsic effective connectivity.

| Analysis | Connections (n) |  |  | Direction of effect | Inhibitory / Excitatory by direction |
| --- | --- | --- | --- | --- | --- |
|  | Total | Inhibitory | Excitatory |  |  |
| Group comparison |  |  |  |  |  |
| PRE > POST | 15 | 9 | 6 | Enhanced in PRE: 7<br>Reduced in PRE: 8 | Enhanced: 2 inh. / 5 exc.<br>Reduced: 7 inh. / 1 exc. |
| PRE > PERI | 19 | 11 | 8 | Enhanced in PRE: 7<br>Reduced in PRE: 12 | Enhanced: 3 inh. / 4 exc.<br>Reduced: 8 inh. / 4 exc. |
| PERI > POST | 11 | 8 | 3 | Enhanced in PERI: 5<br>Reduced in PERI: 6 | Enhanced: 3 inh. / 2 exc.<br>Reduced: 4 inh. / 2 exc. |
| Associations with self-reported emotion regulation ability (DERS-16) |  |  |  |  |  |
| PRE | 12 | 8 | 4 | Positive: 7<br>Negative: 5 | Positive: 4 inh. / 3 exc.<br>Negative: 4 inh. / 1 exc. |
| PERI | 8 | 5 | 3 | Positive: 3<br>Negative: 5 | Positive: 2 inh. / 1 exc.<br>Negative: 3 inh. / 2 exc. |
| POST | 15 | 8 | 7 | Positive: 10<br>Negative: 5 | Positive: 6 inh. / 4 exc.<br>Negative: 2 inh. / 3 exc. |

**Table 3.**
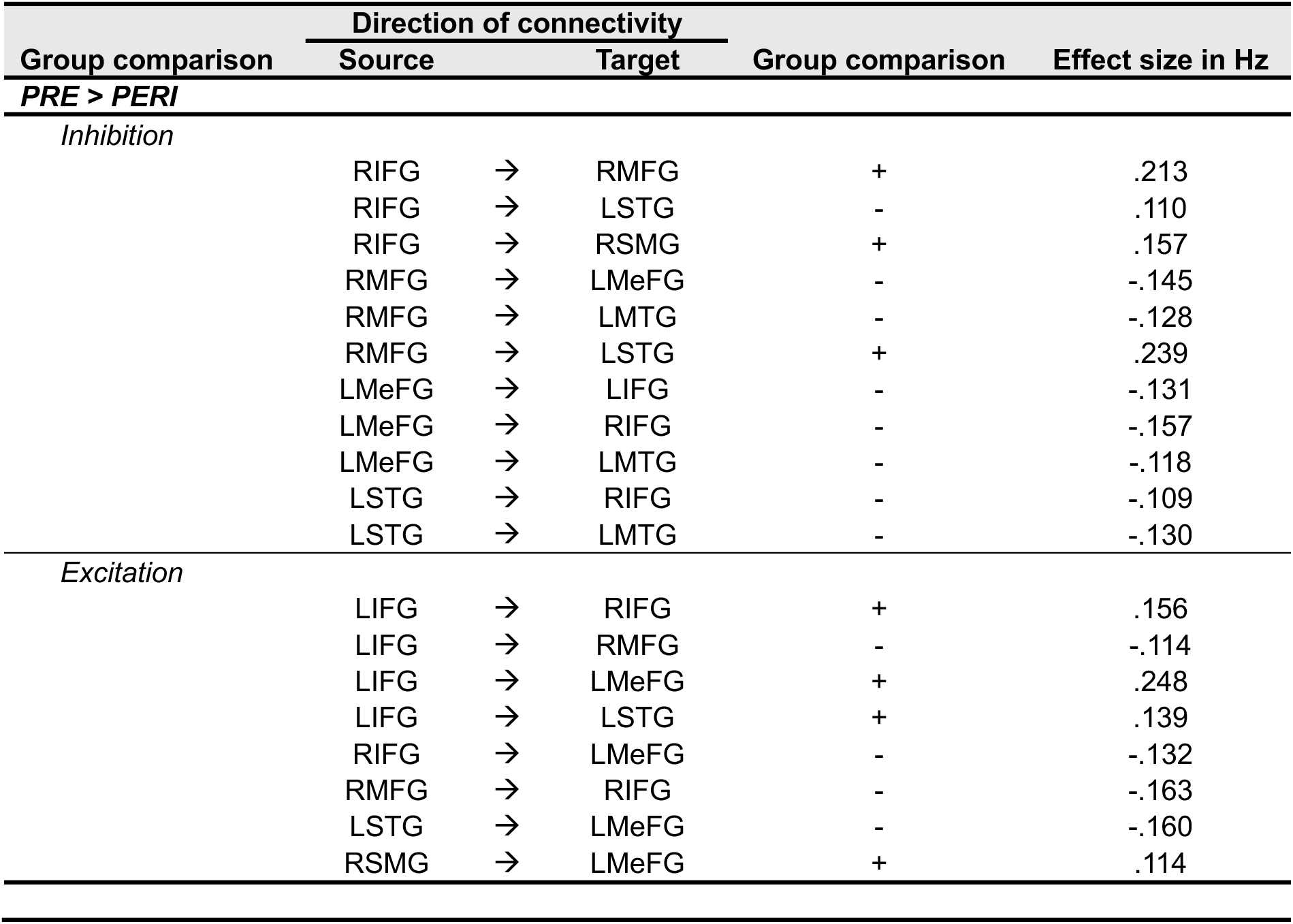
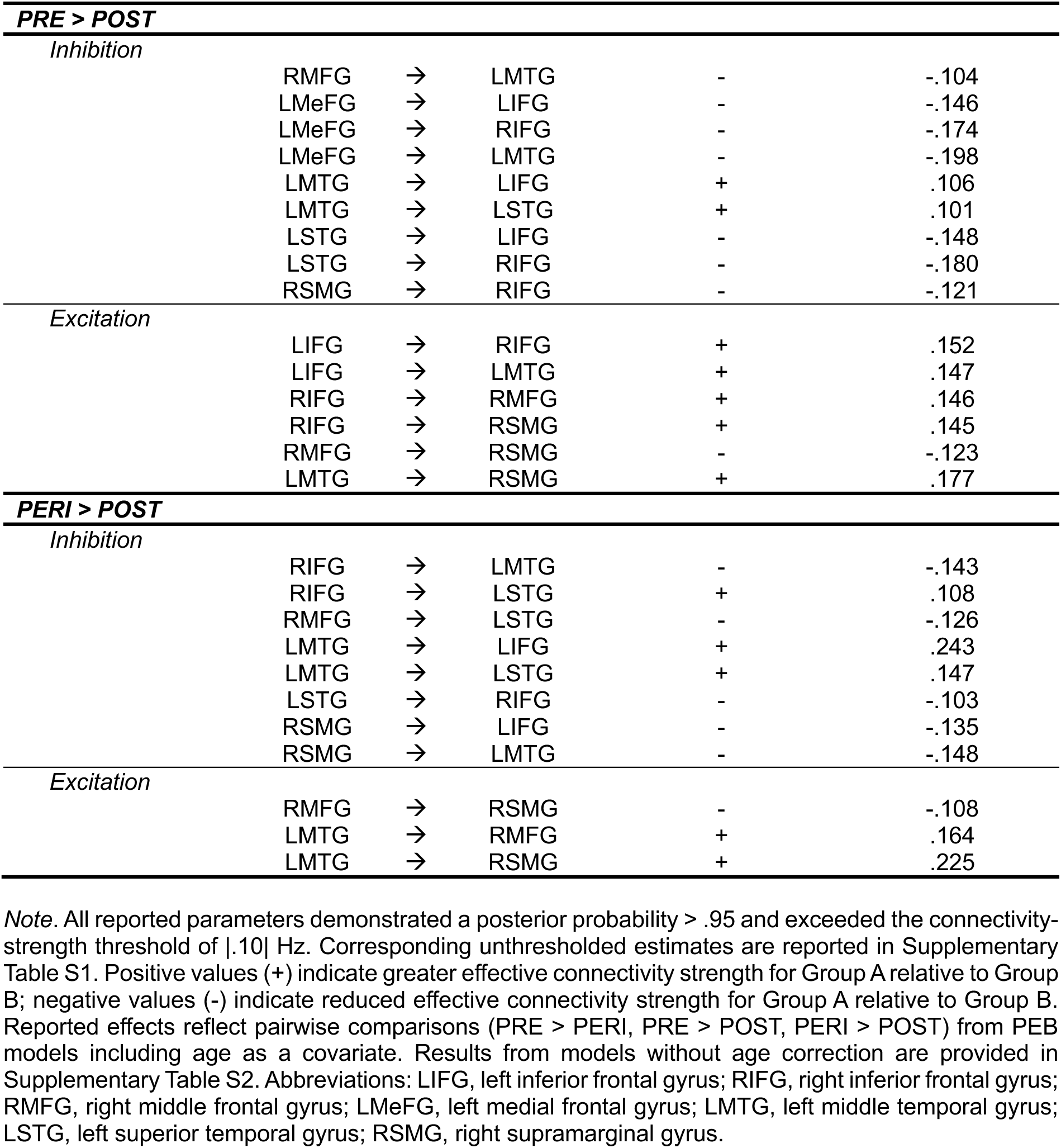
Connections with strong evidence (posterior probability > .95) for group differences in effective connectivity, presenting connections with an absolute connectivity strength ≥ |.10| Hz.

The **PRE vs POST** contrast (Figure 2A) revealed a selective reorganization of effective connectivity within the ER network, characterized by a shift in the balance between excitatory and inhibitory influences (Table 2). Overall, 15 connections differed, with the PRE group showing partially enhanced but predominantly reduced inhibitory connectivity. Specifically, outgoing connectivity in the PRE relative to POST group was exclusively enhanced from bilateral IFG and LMTG and exclusively reduced from the remaining regions of the network (RMFG, LMeFG, LSTG, RSMG). All enhanced connectivity from the IFG was excitatory and targeted frontal, temporal, and parietal regions. The LMTG showed enhanced outgoing connectivity to three regions of the network, comprising inhibitory projections to LIFG and LSTG and excitatory projections to RSMG. Reduced connectivity in PRE compared to POST predominantly reflected inhibitory connections. As a major target region, the RIFG received both enhanced excitatory (from LIFG) and reduced inhibitory inputs (from LMeFG, LSTG, RSMG) in PRE compared to POST. Overall, the PRE state was characterized by a highly focal enhancement of excitatory output from bilateral IFG and LMTG alongside widespread reduction of predominantly inhibitory connections from the rest of the network compared to POST. This resulted in a network configuration marked by increased excitatory drive and relative disinhibition of the RIFG as a central regulatory hub.

The **PRE vs PERI** contrast (Figure 2B) revealed the most extensive group differences in effective connectivity within the ER network, marked by a highly heterogeneous pattern of connectivity differences (Table 2). Overall, 19 connections differed, with PRE showing both enhanced and predominantly reduced connectivity across frontal and temporal pathways. Connectivity differences were dominated by frontal source regions. These regions exhibited mixed profiles of enhanced and reduced projections involving both excitatory and inhibitory connections targeting the rest of the network. Specifically, bilateral IFG and RMFG showed different connectivity patterns targeting frontal, temporal, and parietal regions. In contrast, connectivity originating from the LMeFG and LSTG was exclusively reduced in the PRE compared to PERI group, primarily reflecting inhibitory connections to the IFG and LMTG. As a target hub, the LMeFG showed both increased and decreased incoming connections in PRE compared to PERI, receiving altered inputs from all other regions of the network except the LMTG, with a predominance of excitatory connections. Altogether, connectivity differences between PRE and PERI were widespread and heterogeneous, dominated by frontal regions and characterized by a distinct, mostly excitatory convergence on the LMeFG that was not observed in the other group comparisons.

The **PERI vs POST** contrast (Figure 2C) revealed fewer differences and an absence of frontal-to-frontal connectivity differences within the network. Overall, 11 connections differed, with PERI showing balanced enhanced and reduced connectivity, predominantly involving inhibitory connections (Table 2). Reduced connectivity in PERI relative to POST originated from right frontal regions (RIFG, RMFG) and targeted temporal regions (LMTG, LSTG) via inhibitory connections as well as the parietal region (RSMG) via excitatory connections. In the PERI group, the RSMG showed reduced inhibitory connectivity toward the LIFG and LMTG. Notably, the LMTG emerged as a key source hub, revealing four exclusively enhanced outgoing connections for PERI relative to POST. These included inhibitory connections to LIFG and LSTG, and excitatory connections to RMFG and RSMG. Overall, PERI and POST showed distinct connectivity differences, with reduced emphasis on frontal connections and greater temporo-parietal source involvement, driven primarily by enhanced LMTG output in PERI.

##### Summary across pairwise contrasts

In a descriptive comparison across pairwise contrasts, differences in ER network connectivity varied substantially in both the number and direction of affected connection. Qualitatively, PRE differed most strongly in frontal connectivity, with the PRE vs PERI contrast showing the greatest number and heterogeneity of directed effects. In contrast, PRE vs POST differences were more focal, and PERI vs POST differences were comparatively subtle. Two regions consistently emerged as transition-sensitive hubs: the LMTG acted as a key source hub in contrasts involving POST (PRE vs POST and PERI vs POST), whereas the LMeFG served as a selective target hub only in the PRE vs PERI contrast, receiving convergent inputs which were not observed in contrasts involving POST.

#### Association between self-reported ER ability and effective connectivity in the regulation network

Within-group PEB analyses examined associations between effective connectivity and self-reported ER ability (DERS-16). A concise overview of connectivity–ER associations within each group is provided in Table 3, with thresholded estimates reported in Table 4 and visualized in Figure 3A. Positive values (green) indicate a positive association between effective connectivity and DERS-16 scores, whereas negative values (orange/red) indicate a negative association. Connections marked by asterisks in Figure 3A indicate associations unique to the respective menopausal group (i.e., not observed in the other groups). For interpretability, we report and visualize only connections with an absolute connectivity strength ≥ |.01|Hz, as associations with self-reported ER tended to occur at lower connectivity strengths. Unthresholded results are provided in Supplementary Table S3 and Supplementary Figure S5.

**Figure 3.**
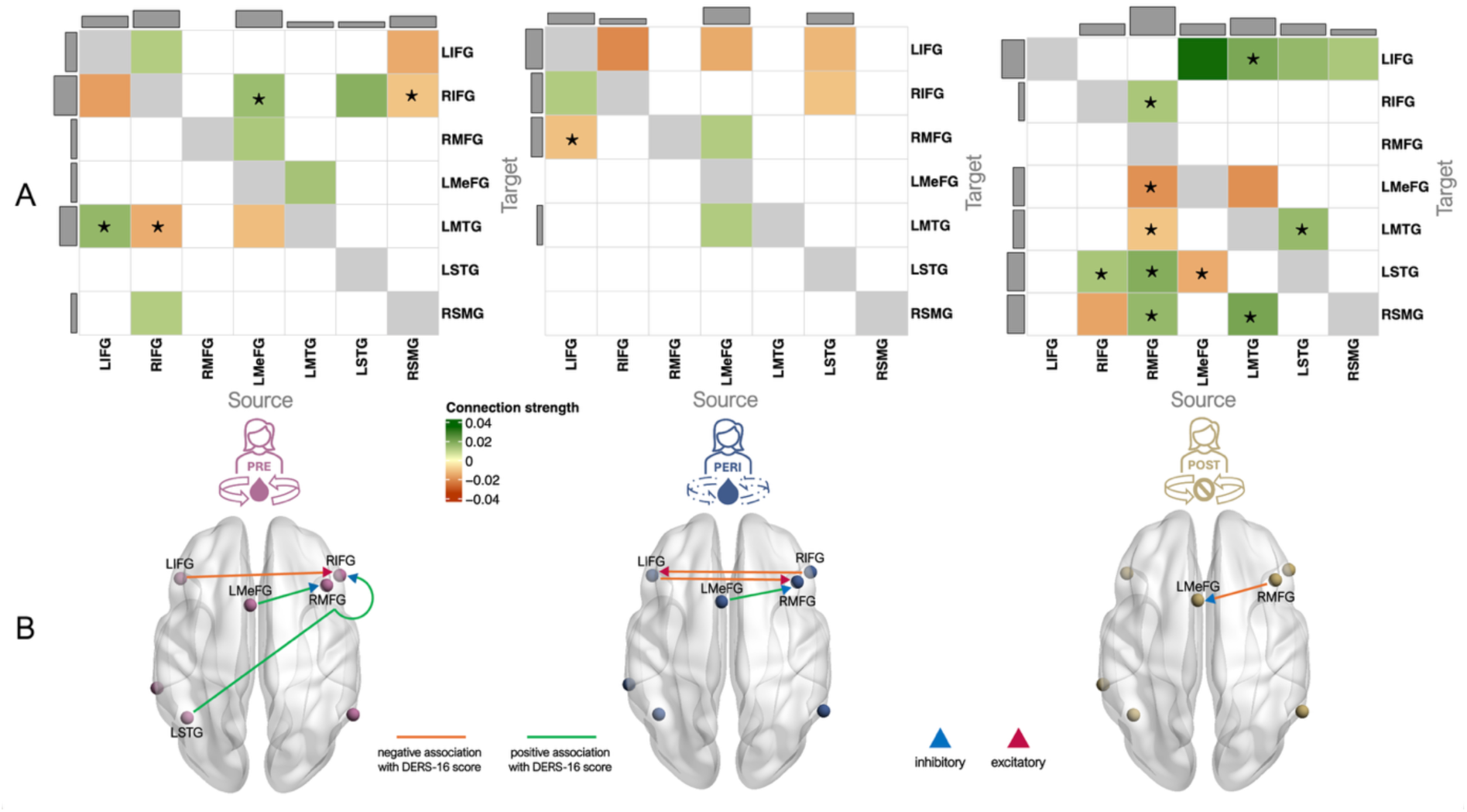
Associations between effective connectivity and self-reported ability in emotion regulation (Difficulties in Emotion Regulation Scale-16, DERS-16). (A) Connections showing strong evidence (posterior probability > 0.95) for associations with DERS-16 scores within each group; only connections with an absolute connectivity strength ≥ |.01| Hz are shown. Asterisks indicate connections uniquely associated with DERS-16 within a given group (i.e., not observed in the other groups). (B) Leave-one-out cross-validation (LOOCV) results assessing the predictive validity of DERS-16 scores. *Note*. Displayed connections in (A) met both the posterior-probability criterion (PP > .95) and the absolute connectivity-strength threshold (|.01| Hz). Positive values (green) indicate a positive association between effective connectivity and DERS-16 scores, whereas negative values (orange/red) indicate a negative association in (A) and (B). Arrow-head color in (B) indicates excitatory (red) and inhibitory (blue) connectivity. Abbreviations: LIFG, left inferior frontal gyrus; RIFG, right inferior frontal gyrus; RMFG, right middle frontal gyrus; LMeFG, left medial frontal gyrus; LMTG, left middle temporal gyrus; LSTG, left superior temporal gyrus; RSMG, right supramarginal gyrus.

**Table 4.**
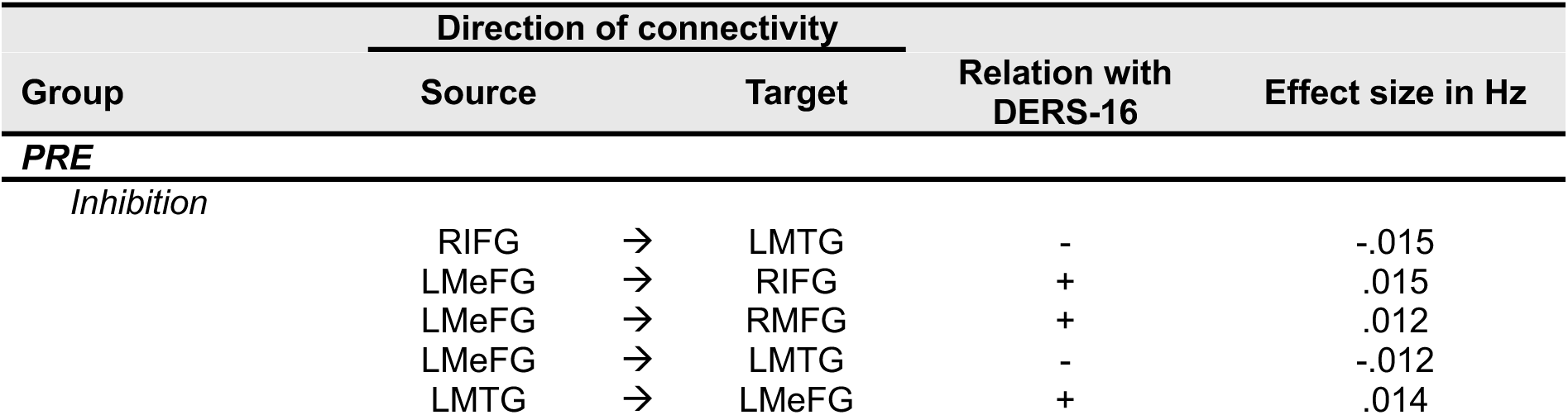
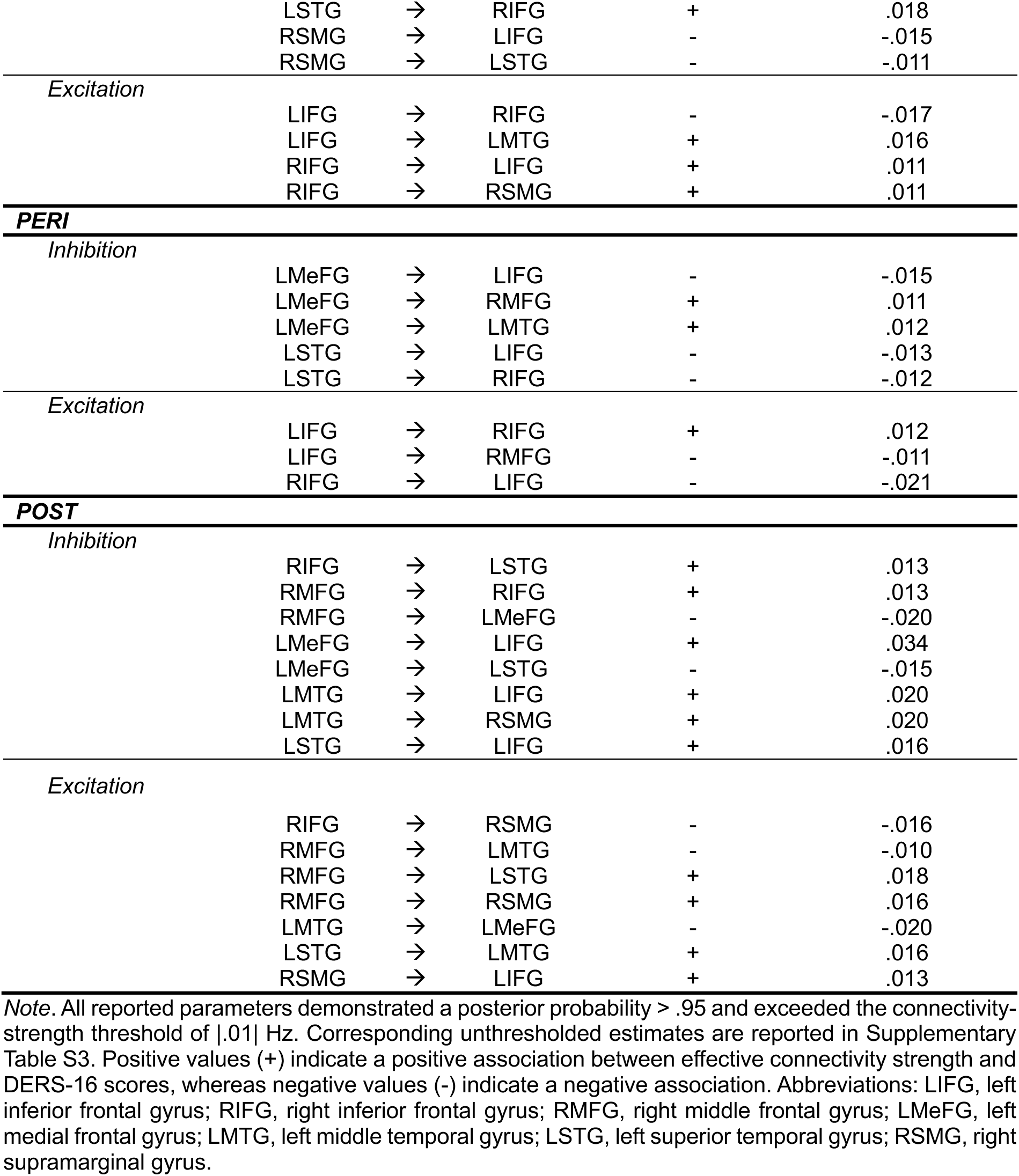
Connections with strong evidence (posterior probability > .95) for associations between effective connectivity and self-reported emotion regulation ability (DERS-16), presenting connections with an absolute connectivity strength ≥ |.01| Hz.

In the **PRE** group, ER ability was associated with a distributed pattern of connectivity involving both inhibitory and excitatory influences across frontal, temporal, and parietal regions (Figure 3A). The RIFG emerged as a central hub, acting both as a major target and, together with the LMeFG, as a primary source of ER-related connectivity. Frontal regions exhibited a mixed profile of positive and negative associations, with bilateral IFG contributing predominantly excitatory connections and the LMeFG exclusively inhibitory connections. Temporal-to-frontal connections were consistently positively associated with DERS-16, whereas the RSMG contributed inhibitory projections to bilateral IFG that were negatively associated with DERS-16.

In the **PERI** group, ER ability-related connectivity was largely restricted to frontal regions and characterized by a predominance of negative associations (Figure 3A). The LMeFG emerged as the principal source hub, contributing exclusively inhibitory projections with differential associations to DERS-16, including negative associations to the LIFG and positive associations to the RMFG and LMTG. The LIFG acted as the primary target hub, receiving exclusively negatively associated connections, including inhibitory projections from the LMeFG and LSTG and excitatory projections from the RIFG. The LSTG was the only temporal region involved, contributing inhibitory connections to bilateral IFG which were negatively associated with DERS-16.

In the **POST** group, ER ability was associated with a more extensive and interconnected network pattern (see Figure 3A). The RMFG emerged as a prominent source hub, connecting to frontal, temporal, and parietal regions with heterogeneous associations. RMFG-derived connections showed positive associations to the RIFG, LSTG, and RSMG, alongside negative associations to the LMeFG and LMTG. In contrast, the LIFG acted as the principal target hub, receiving exclusively positively associated connections. Inputs to the LIFG from frontal (LMeFG) and temporal (LMTG, LSTG) sources were inhibitory, whereas the parietal input (RSMG) was excitatory.

##### Summary across groups

In a qualitative comparison across groups, connectivity patterns associated with self-reported ER ability differed markedly in extent, directionality, and network organization. PRE showed a distributed association pattern; PERI exhibited a restricted, frontal-focused pattern; and POST showed the most extensive associations, reflecting broader network-interconnectedness. Notably, the RMFG emerged as POST-specific source hub, whereas the LIFG showed a stage-dependent target profile, showing mixed associations in PRE, negative associations in PERI, and exclusively positive associations in POST.

#### Prediction of self-report ER ability

Finally, we assessed whether DERS-16 scores could be predicted from effective connectivity within the regulation network using LOOCV. Only connections exceeding a posterior probability > .95 in the association models were entered into the prediction analysis. Results are shown in Figure 3B and reported in Table 5. In the **PRE** group, three connections predicted DERS-16 scores: while the excitatory projection from LIFG to RIFG showed a negative association, two inhibitory projections, from LMeFG to RMFG and from LSTG to RIFG, showed positive associations. Similarly, in the **PERI** group, three predictive frontal connections were found: Connections from the LIFG to RMFG and from the RIFG to LIFG were excitatory and negatively associated with DERS-16 scores, whereas the connection from LMeFG to RMFG was inhibitory and positively associated, thus, mirroring the predictive role of this connection observed in the PRE group. In contrast, only a single inhibitory connection from the RMFG to LMeFG was significant in the **POST** group, indicating a negative prediction of DERS-16 scores.

**Table 5.**
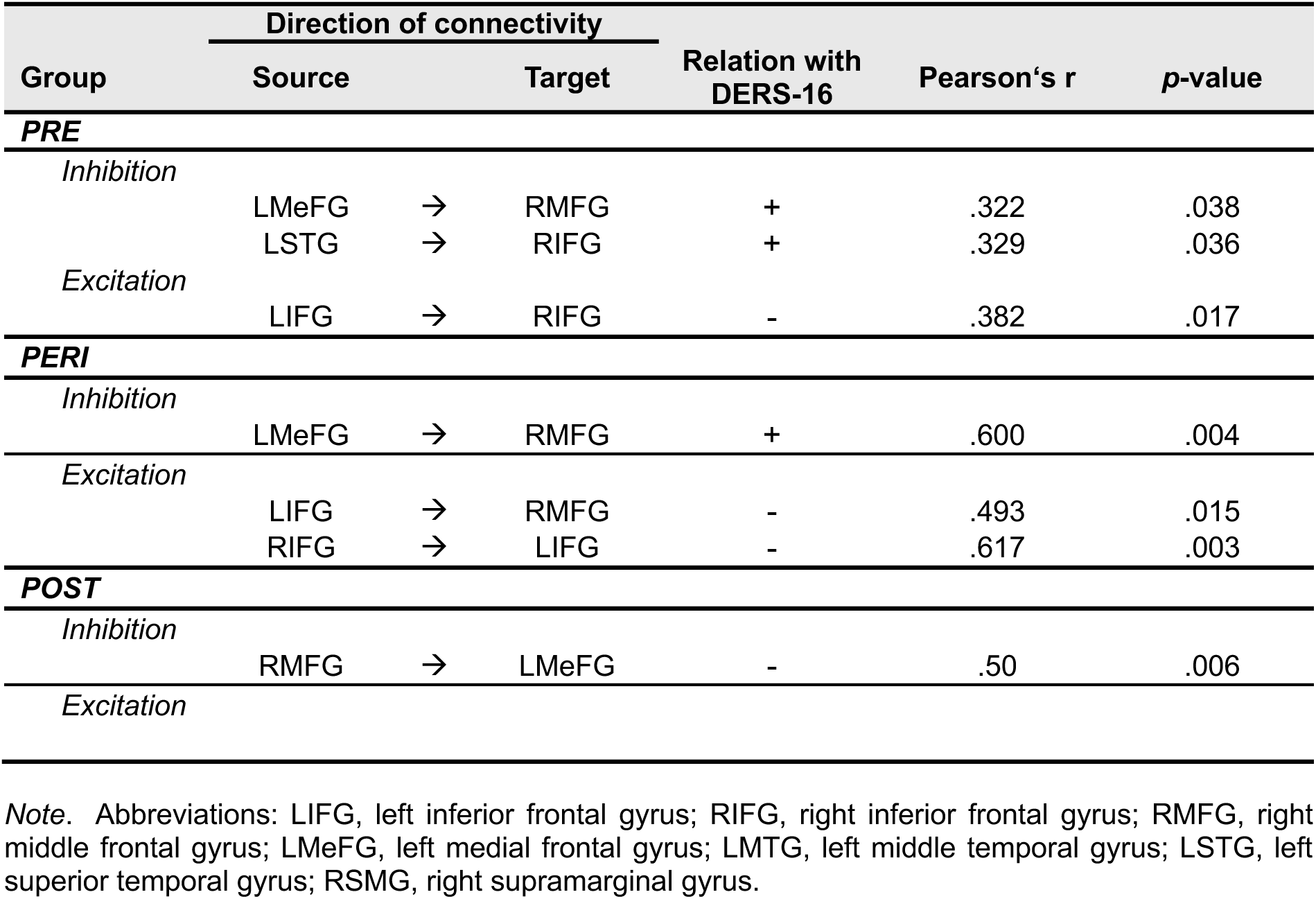
Leave-one-out cross-validation (LOOCV) results for predicting self-reported emotion regulation ability (Difficulties in Emotion Regulation Scale-16, DERS-16).

| Group | Direction of connectivity |  |  | Relation with DERS-16 | Pearson's r | p-value |
| --- | --- | --- | --- | --- | --- | --- |
|  | Source |  | Target |  |  |  |
| PRE |  |  |  |  |  |  |
| Inhibition |  |  |  |  |  |  |
|  | LMeFG | → | RMFG | + | .322 | .038 |
|  | LSTG | → | RIFG | + | .329 | .036 |
| Excitation |  |  |  |  |  |  |
|  | LIFG | → | RIFG | - | .382 | .017 |
| PERI |  |  |  |  |  |  |
| Inhibition |  |  |  |  |  |  |
|  | LMeFG | → | RMFG | + | .600 | .004 |
| Excitation |  |  |  |  |  |  |
|  | LIFG | → | RMFG | - | .493 | .015 |
|  | RIFG | → | LIFG | - | .617 | .003 |
| POST |  |  |  |  |  |  |
| Inhibition |  |  |  |  |  |  |
|  | RMFG | → | LMeFG | - | .50 | .006 |
| Excitation |  |  |  |  |  |  |

*Note.* Abbreviations: LIFG, left inferior frontal gyrus; RIFG, right inferior frontal gyrus; RMFG, right middle frontal gyrus; LMeFG, left medial frontal gyrus; LMTG, left middle temporal gyrus; LSTG, left superior temporal gyrus; RSMG, right supramarginal gyrus.

## Discussion

Menopause is a major psychoneuroendocrine transition which can impact emotional functioning and mental health [6–9]. Although ER is fundamental for mental health [31,34], intrinsic neural connectivity supporting ER during the menopausal transition has remained unexplored. Addressing this gap, the present study examined directed resting-state effective connectivity within a predefined ER network [64] across healthy pre-, peri-, and postmenopausal women using spDCM [53]. We demonstrate menopausal stage-specific differences in intrinsic ER network connectivity, suggesting a redistribution of network organization. Particularly, perimenopause emerged as a distinct phase characterized by widespread and heterogeneous connectivity patterns not evident in pre-versus postmenopausal comparisons alone. Importantly, these intrinsic neural differences emerged in the absence of group differences in self-reported ER ability, suggesting preserved trait-based ER ability despite distinct baseline network architectures before, during, and after menopause.

### Perimenopause as a distinct phase of intrinsic ER network organization rather than a transitional midpoint

Many neuroimaging studies on the menopausal transition have relied on cross-sectional contrasts between two stages, most commonly pre-vs postmenopause [88–91], and less frequently pre-vs perimenopause [92,93] or peri-vs postmenopause [94]. Other studies combine pre- and perimenopausal women for comparison with postmenopause [95,96]. This literature, predominately based on binary group contrasts, is commonly interpreted within a graded, stepwise framework. When considered in isolation, our PRE vs POST comparison likewise showed clear differences in intrinsic ER network connectivity, characterized by frontal alongside a set of temporal connectivity differences. Yet the full pattern of network reorganization across the menopausal transition only becomes visible when perimenopause is modeled as a distinct transition stage. PRE vs PERI showed the greatest number and heterogeneity of connectivity differences, particularly pronounced for frontal connections. PERI vs POST showed fewer differences and a qualitatively shifted profile toward temporal and parietal sources. This non-monotonic pattern, rather than simple intermediate between pre-and postmenopause, aligns with resting-state evidence for perimenopause-specific alterations in intrinsic function relative to both premenopause [93] and postmenopause [94]. Consistent findings from structural, metabolic, and task-based fMRI studies further identify perimenopause as a distinct neural phase [14,26,97–99]. More broadly, spDCM work demonstrates that intrinsic connectivity is sensitive to endogenous hormonal variation in a stage-dependent manner across the menstrual cycle, as shown in within-subject designs [54], and that estradiol modulates effective connectivity within ER networks at rest [52]. Given the pronounced endocrine fluctuations that characterize the menopausal transition [17], hormone-sensitive models provide a relevant framework for examining intrinsic effective connectivity changes also during the menopausal transition. Our findings provide first evidence that intrinsic ER network organization undergoes dynamic, stage-specific differences across the menopausal transition, with perimenopause presenting a distinct and critical window of neural reorganization [8,26].

#### Transition-sensitive hubs within the ER network

To elucidate stage-specific differences in intrinsic ER network organization, we focus on three transition-sensitive hubs within the ER network: the bilateral IFG, LMeFG, and LMTG. Connectivity patterns of these regions varied across menopausal stages. In PRE, the IFG showed strong network engagement, characterized by enhanced excitatory output and reduced inhibitory input relative to POST. This pattern is consistent with task-based effective connectivity findings in younger samples that position lateral prefrontal regions as central nodes for ER selection and implementation via balanced frontal interactions [42,43]. Within a resting-state framework, this IFG-dominant profile may indicate a preparatory control configuration relevant for ER when required [63]. Given evidence that estradiol modulates prefrontal inhibitory connectivity within the ER network [52], such a preparatory configuration in premenopause could be supported by relatively higher estradiol levels compared with the fluctuating and declining levels observed in peri- and postmenopause [17]. In contrast, the PRE vs PERI comparison revealed a more heterogeneous IFG profile, with divergent outgoing connections to frontal regions, consistent with resting-state evidence for altered intrinsic frontal organization during perimenopause [93,94]. The absence of frontal–frontal differences in the PERI vs POST contrast suggests that the frontal connectivity pattern is most pronounced around the transition into perimenopause and subsequently stabilizes or shifts toward other network components. The LMeFG showed a complementary stage-dependent profile. As a source region, the LMeFG had reduced inhibitory output in PRE relative to both PERI and POST, with no differences between the latter two groups. As a target, the LMeFG emerged as a transient convergence hub uniquely in the PRE > PERI contrast, receiving heterogeneous excitatory inputs from nearly all other regions. This pattern emphasizes a temporary reweighting of frontal intrinsic network organization around perimenopause, rather than a monotonic shift across stages. Given the role of the LMeFG in coordinating information flow among lateral frontal regions during ER [43,100], our findings could point to differences in baseline intrinsic frontal control organization across menopausal stages. Beyond frontal regions, the LMTG emerged as a key temporal source hub in contrasts involving POST. Both PRE and PERI showed enhanced LMTG output to frontal, temporal, and parietal targets relative to POST, with selective enhancement of LMTG–frontal connectivity in PERI. This pattern further supports stage-specific differences in how temporal regions are intrinsically coupled within the ER network, rather than a graded progression across stages. While the LMTG is consistently implicated in ER-related processing in task-related studies [38,43,63,101], the present findings indicate that its intrinsic coupling with frontal regions differs across menopausal stages, highlighting non-uniform baseline ER network architecture. Future longitudinal studies are needed to confirm these stage-specific connectivity patterns and their endocrine modulation. Critically, across these transition-sensitive hubs, the observed patterns reflect differences in baseline, preparatory network organization rather than direct evidence for altered ER performance [63], particularly considering preserved trait-based ER ability.

### Preserved subjective ER ability alongside stage-specific intrinsic network organization

Despite pronounced stage-specific differences in intrinsic ER network connectivity, groups did not differ in self-reported ER ability. This dissociation indicates that comparable levels of perceived ER ability can be maintained across the menopausal transition alongside qualitative differences in intrinsic network configuration. Similar dissociations between neural measures and ER outcomes have been reported in younger and premenopausal samples, where hormone-related modulation of brain–behavior relationships occurred in the absence of differences in overt ER performance [51,52], and are consistent with models emphasizing flexibility in intrinsic ER network organization [63]. In the present sample, perimenopausal women reported higher negative affect than premenopausal women despite comparable self-reported ER ability, indicating that changes in affective experience can emerge independently of perceived regulatory ability. While affective states can modulate functional connectivity, particularly under experimental manipulation [102,103], resting-state effective connectivity is generally interpreted as reflecting a relatively stable, trait-like baseline organization [63]. Accordingly, the observed stage-specific connectivity patterns are unlikely to be driven by transient affect alone.

Associations between intrinsic connectivity and trait-based ER ability differed across menopausal stages with respect to regional involvement, directionality, and predictive validity. Analyses converged with group-level effects and identified the transition-sensitive hubs, particularly IFG and LMeFG, as key regions linking intrinsic connectivity to self-reported ER ability. In PRE, ER ability was associated with a distributed connectivity profile spanning frontal and temporal regions, in which IFG and LMeFG emerged as primary hubs. This is consistent with prior ER network studies in younger samples showing multi-node involvement [36,38,42,43,63]. PERI showed a restricted, frontal-focused profile, characterized by LMeFG-source and LIFG-target connectivity, suggesting that variability in trait-based ER ability during perimenopause is captured by a narrower subset of intrinsic frontal interactions. POST exhibited further qualitative differences, with ER-related associations involving LIFG targets and LMTG sources. The single frontal connection from the RMFG to LMeFG which was predictive of self-reported ER ability in POST showed reversed directionality relative to PRE and PERI. This suggests a qualitative change in how intrinsic connectivity relates to trait-based ER after the transition and supports evidence for postmenopausal neural adaptation [26]. Within a resting-state framework, these findings support IFG, LMeFG, and LMTG as transition-sensitive hubs whose connectivity profiles, targets, and directionality vary in a stage-dependent manner. The coexistence of preserved self-reported ER ability with distinct intrinsic connectivity patterns suggests that similar levels of trait-based ER may be supported by functionally equivalent, but qualitatively different, baseline network configurations across menopausal stages. In this context, perimenopause emerges as a differentiated intrinsic network state rather than a graded intermediate between pre- and postmenopause, reflecting stage-specific organization of the ER network at rest, in anticipation of emotional demands.

### Limitations, strengths, and future directions

Several limitations warrant consideration. First, the cross-sectional design precludes inferences about within-individual trajectories across the menopausal transition. Nonetheless, this study provides the first application of spDCM to intrinsic effective connectivity across distinct menopausal stages while explicitly modeling perimenopause as a unique neuroendocrine transition window. By modeling directed connectivity rather than undirected functional coupling, this approach reveals stage-specific network organization that may not be detectable using conventional resting-state analyses. Longitudinal studies in larger cohorts are needed to characterize temporal trajectories and interindividual variability in intrinsic network adaptation. Second, hormone levels and menopausal symptoms were not modeled explicitly, and a small subset of peri- and postmenopausal participants reported current MHT use. Although ovarian hormones influence intrinsic connectivity [52,54,104], and menopausal symptoms have been linked to resting-state activity [105,106] and ER difficulties [60], the present study focused on menopausal stage as a relevant proxy. Importantly, PERI and POST groups did not differ in symptom severity, reducing the likelihood that symptom burden alone accounted for the observed effects. As prior findings on MHT-related neural effects remain mixed [27,107], future work should incorporate stratified or interventional designs to understand how hormonal variability and MHT shape intrinsic network organization across the menopausal transition. Third, the use of resting-state data indexes baseline network organization and limits conclusions about task-evoked ER processes. Additionally, while anchoring analyses in a predefined ER network enabled integration with prior intrinsic connectivity work [52], effective ER depends on dynamic interactions across multiple large-scale networks and context-dependent processes [38,63]. Future studies should extend this approach by examining additional ER-relevant networks and between-network connectivity, ideally combining intrinsic and task-based effective connectivity to directly link baseline network configuration to regulatory strategies and behavioral outcomes. Fourth, effective connectivity may be influenced by factors inherent to resting-state acquisitions, including vigilance, arousal, or sensory input. To mitigate this, participants viewed a low-demand, non-narrative video (Inscapes), which reduces head motion and sleep while preserving resting-state conditions [75]. While this approach improves stability and comparability of baseline connectivity across groups, it may limit direct comparability with resting-state studies using eyes-open or eyes-closed conditions. Finally, the present findings are derived from a mentally healthy sample, thus, primarily inform intrinsic network organization under conditions of preserved emotional functioning, with limited variability in ER ability potentially constraining detectable group differences in self-reports. Extending this work to clinical populations will be essential to determine whether intrinsic ER network organization may differ during the menopausal transition [15]. Together, these considerations highlight the need for longitudinal, multimodal approaches integrating intrinsic and task-based connectivity, hormonal dynamics, and clinical outcomes to determine when stage-specific intrinsic network configurations support adaptive regulation and when they may confer affective risk.

## Conclusion

This study provides first evidence that intrinsic ER network connectivity differs in a non-monotonic, stage-dependent pattern across the menopausal transition. Despite preserved trait-based ER ability, resting-state effective connectivity within a predefined ER network varied qualitatively between pre-, peri-, and postmenopause, indicating that baseline network organization is not uniform across stages. Notably, perimenopause showed a distinct pattern of intrinsic network connectivity instead of a graded intermediate midpoint between pre- and postmenopause, highlighting it as a unique neuroendocrine transition period. These findings extend hormone-sensitive models of intrinsic connectivity by demonstrating that psychoneuroendocrine transitions are associated with qualitative differences in baseline ER network configuration in healthy women. More broadly, the present findings provide a foundation for future longitudinal and multimodal research aimed at prospectively linking intrinsic network organization, hormonal dynamics, and ER as well as clinical outcomes. Such approaches will be essential to determine when stage-specific intrinsic network configurations reflect resilience and when they may confer vulnerability, thereby informing targeted strategies to promote emotional well-being across the menopausal transition and beyond.

## Glossary

AMH: Anti-Müllerian hormone
BMA: Bayesian model averaging
BMR: Bayesian model reduction
BOLD: Blood-oxygenation-level-dependent
DCM: dynamic causal modeling
spDCM: spectral dynamic causal modeling
DERS-16: Difficulties in Emotion Regulation Scale-16
EPIs: Echo-planar images
ER: emotion regulation
FSH: follicle-stimulating hormone
LH: luteinizing hormone
LIFG: left inferior frontal gyrus
LMeFG: left medial frontal gyrus
LMTG: left middle temporal gyrus
LOOCV: leave-one-out cross-validation
LSTG: left superior temporal gyrus
MRI: magnetic resonance imaging
fMRI: functional magnetic resonance imaging
rs-fMRI: resting-state functional magnetic resonance imaging
PEB: parametric empirical Bayes
PERI: perimenopausal women
PFC: prefrontal cortex
POST: postmenopausal women
PRE: premenopausal women
RIFG: right inferior frontal gyrus
RMFG: right middle frontal gyrus
ROI: region-of-interest
RSMG: right supramarginal gyrus
SHBG: sex hormone-binding globulin

## Acknowledgements

We would like to thank all the women who participated in this study for their time and effort. We acknowledge Luise Pfleiderer, Katharina Stein, Julia Graf, and Corinna Stramazzo for their assistance in data collection. We further acknowledge the Core Facility of the University of Tübingen for providing access to MRI infrastructure and for their technical support during data collection.

## Author Contribution

FW: Conceptualization, Data curation, Formal analysis, Investigation, Methodology, Project administration, Software, Validation, Visualization, Writing – original draft. ACSK: Data curation, Project administration, Software, Supervision, Validation, Writing – review and editing. SA: Formal analysis, Methodology, Software, Supervision, Validation, Writing – review and editing. LG: Formal analysis, Methodology, Software, Validation, Writing – review and editing. AS: Conceptualization, Funding acquisition, Supervision, Validation, Writing – review and editing. CM: Formal analysis, Methodology, Resources, Software, Supervision, Validation, Writing – review and editing. BD: Conceptualization, Funding acquisition, Project administration, Resources, Supervision, Validation, Writing – review and editing.

## Declaration of AI use

During the preparation of this work the author(s) used ChatGPT (OpenAI, San Francisco, CA) to assist with language editing. After using this tool/service, the author(s) reviewed and edited the content as needed and take(s) full responsibility for the content of the publication.

## Data Availability

The data used in the present study can be requested by sending a research proposal to the principal investigator, Prof. Dr. Birgit Derntl (E-Mail). Please provide a clear and informative title for your proposed research. Please briefly describe the overall rationale for your study and summarize the specific aims/hypotheses that you will test with the specific data elements you are requesting. A data sharing agreement is subsequently drawn up.

