## Supplementary Information for "Distinct intrinsic neural connectivity of an emotion regulation network across the menopausal transition"

### Supplementary Methods

#### Structural control analyses

To assess potential group differences in brain morphology, structural T1-weighted images were preprocessed and segmented using the Computational Anatomy Toolbox (CAT12; https://neuro-jena.github.io/cat//index.html#VBM; Gaser et al., 2024) implemented in SPM12. Individual estimates of total intracranial volume (TIV), gray matter volume (GMV), and white matter volume (WMV) were extracted for each participant. Group differences in global structural measures (TIV, GMV, WMV) were assessed using one-way analyses of variance (ANOVAs) with menopausal status (PRE, PERI, POST) as the between-subjects factor. To account for potential age effects, additional analyses of covariance (ANCOVAs) were conducted including age as a covariate. No significant group differences were observed for any global structural measure (all *p*s ≤ .096), and all effects remained nonsignificant after adjusting for age (all *p*s ≤ .536). Accordingly, global brain volumes were considered comparable across groups and were not included as covariates in subsequent analyses. To further exclude the possibility that group differences in effective connectivity were driven by structural variation within the emotion regulation (ER) network, regional GMV was examined within the network-specific anatomical masks defined by Morawetz et al. (2023). Gray matter images were smoothed with an 8-mm full-width at half-maximum (FWHM) Gaussian kernel and entered into one-way ANOVAs with menopausal status (PRE, PERI, POST) as the between-subjects factor. A small group effect was observed at the unadjusted level ($p$ = .032); however, this effect was fully attenuated when age was included as a covariate ($p$ = .656), indicating no age-independent group differences in region-of-interest-level GMV. Based on these results, gray matter volume was not included as a covariate in the spectral dynamic causal modeling (spDCM) analyses.

**fMRI data preprocessing with *fMRIPrep***

***Preprocessing of $B_0$ inhomogeneity mappings***

A total of 1 fieldmap was found available within the input BIDS structure for this particular subject. A $B_0$ nonuniformity map (or *fieldmap*) was estimated from the phase-drift map(s) measure with two consecutive GRE (gradient-recalled echo) acquisitions. The corresponding phase-map(s) were phase-unwrapped with prelude (FSL None).

***Functional data preprocessing***

For each BOLD run found per subject the following preprocessing was performed. First, a reference volume was generated, using a custom methodology of *fMRIPrep*, for use in head motion correction. Head-motion parameters with respect to the BOLD reference (transformation matrices, and six corresponding rotation and translation parameters) are estimated before any spatiotemporal filtering using mcflirt (FSL, Jenkinson et al., 2002). The estimated *fieldmap* was then aligned with rigid-registration to the target EPI (echo-planar imaging) reference run. The field coefficients were mapped on to the reference EPI using the transform. The BOLD reference was then co-registered to the T1w reference using bbregister (FreeSurfer) which implements boundary-based registration (Greve & Fischl, 2009). Co-registration was configured with six degrees of freedom. The aligned T2w image was used for initial co-registration. Several confounding time-series were calculated based on the *preprocessed BOLD*: framewise displacement (FD), DVARS and three region-wise global signals. FD was computed using two formulations following Power (absolute sum of relative motions, Power et al. (2014)) and Jenkinson (relative root mean square displacement between affines, Jenkinson et al. (2002)). FD and DVARS are calculated for each functional run, both using their implementations in *Nipype* (following the definitions by Power et al., 2014). The three global signals are extracted within the CSF, the WM, and the whole-brain masks. Additionally, a set of physiological regressors were extracted to allow for component-based noise correction (*CompCor*, Behzadi et al. 2007). Principal components are estimated after high-pass filtering the *preprocessed BOLD* time-series (using a discrete cosine filter with 128s cut-off) for the two *CompCor* variants: temporal (tCompCor) and anatomical (aCompCor). tCompCor components are then calculated from the top 2% variable voxels within the brain mask. For aCompCor, three probabilistic masks (CSF, WM and combined CSF+WM) are generated in anatomical space. The implementation differs from that of Behzadi et al. in that instead of eroding the masks by 2 pixels on BOLD space, a mask of pixels that likely contain a volume fraction of GM is subtracted from the aCompCor masks. This mask is obtained by dilating a GM mask extracted from the FreeSurfer's *aseg* segmentation, and it ensures components are not extracted from voxels containing a minimal fraction of GM. Finally, these masks are resampled into BOLD space and binarized by thresholding at 0.99 (as in the original implementation). Components are also calculated separately within the WM and CSF masks.

For each CompCor decomposition, the *k* components with the largest singular values are retained, such that the retained components' time series are sufficient to explain 50 percent of variance across the nuisance mask (CSF, WM, combined, or temporal). The remaining components are dropped from consideration. The head-motion estimates calculated in the correction step were also placed within the corresponding confounds file. The confound time series derived from head motion estimates and global signals were expanded with the inclusion of temporal derivatives and quadratic terms for each (Satterthwaite et al. 2013). Frames that exceeded a threshold of 0.5 mm FD or 1.5 standardized DVARS were annotated as motion outliers. Additional nuisance timeseries are calculated by means of principal components analysis of the signal found within a thin band (*crown*) of voxels around the edge of the brain, as proposed by (Patriat et al., 2017). All resamplings can be performed with *a single interpolation step* by composing all the pertinent transformations (i.e. head-motion transform matrices, susceptibility distortion correction when available, and co-registrations to anatomical and output spaces). Gridded (volumetric) resamplings were performed using nitransforms, configured with cubic B-spline interpolation. Many internal operations of *fMRIPrep* use *Nilearn* 0.10.4 (Abraham et al., 2014, RRID:SCR_001362), mostly within the functional processing workflow. For more details of the pipeline, see the section corresponding to workflows in *fMRIPrep*'s documentation.

### Supplementary Results

**Supplementary Figure S1.** Posterior correlations among effective connectivity parameters estimated from spectral DCM for PRE-, PERI-, and POST-menopausal groups.

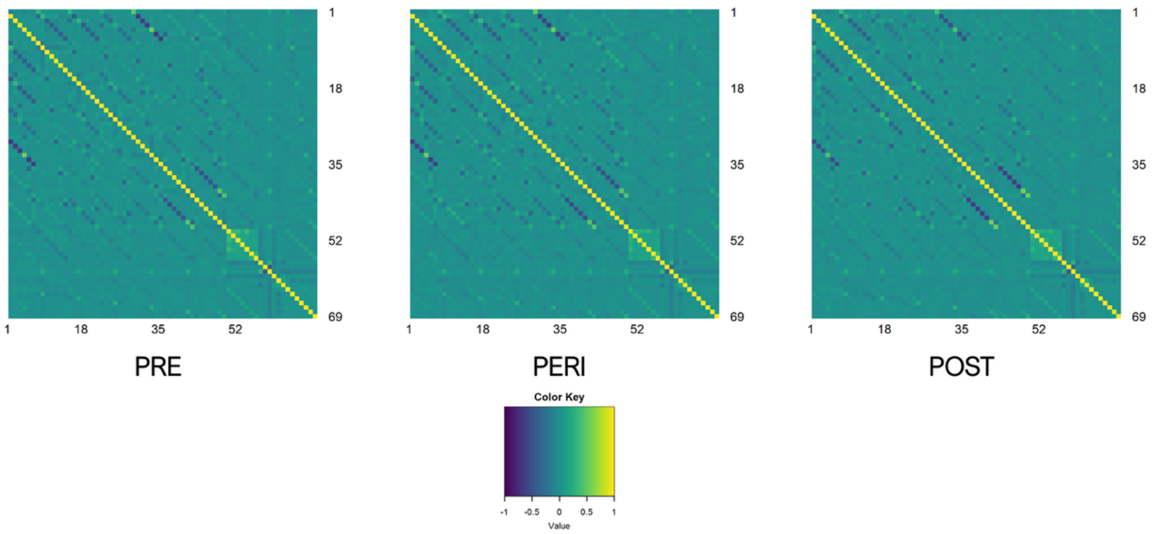

**Supplementary Figure S2. First-level spectral DCM convergence diagnostics for PRE-, PERI-, and POST-menopausal groups:** (A) variance explained for each participant, (B) the largest absolute parameter estimate, and (C) the effective number of parameters, reflecting divergence between posterior and prior densities.

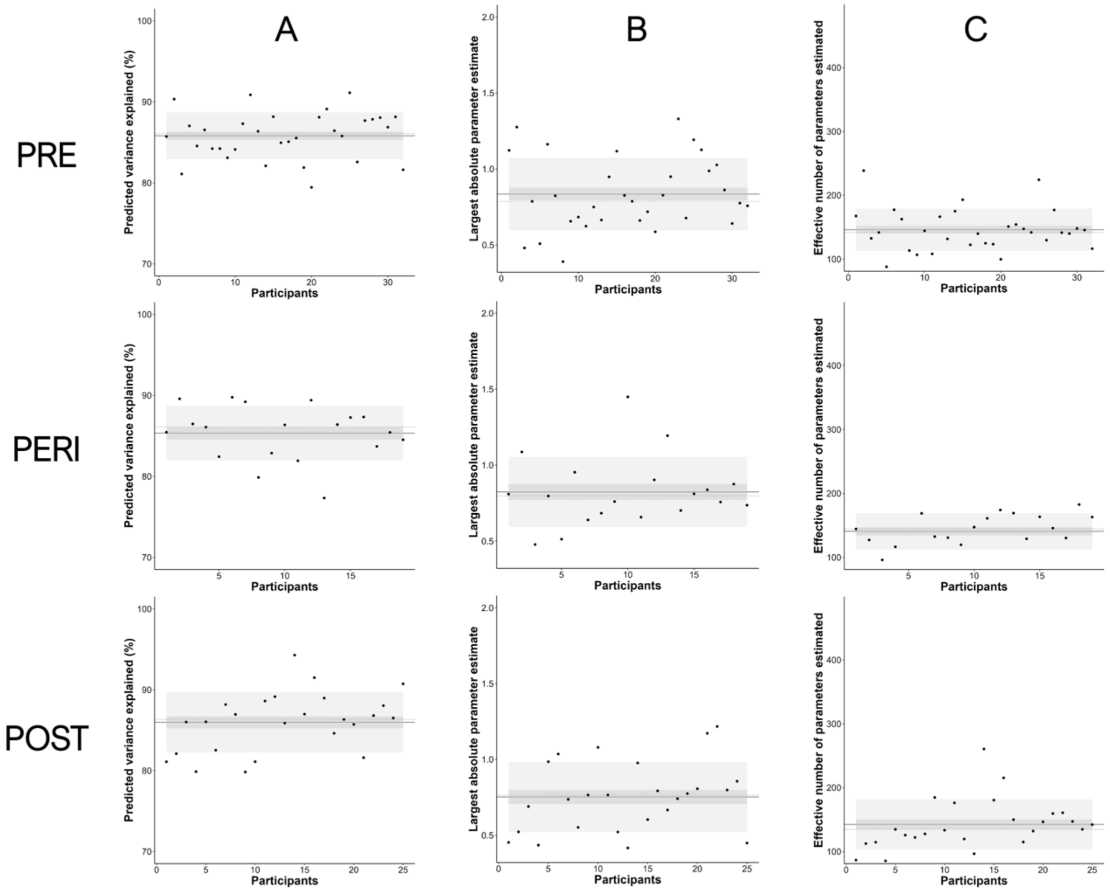

**Supplementary Figure S3. Connections showing strong evidence (posterior probability > .95) for group differences in effective connectivity across menopausal transition without applying a connectivity-strength threshold:** (A) PRE > POST, (B) PRE > PERI, and (C) PERI > POST.

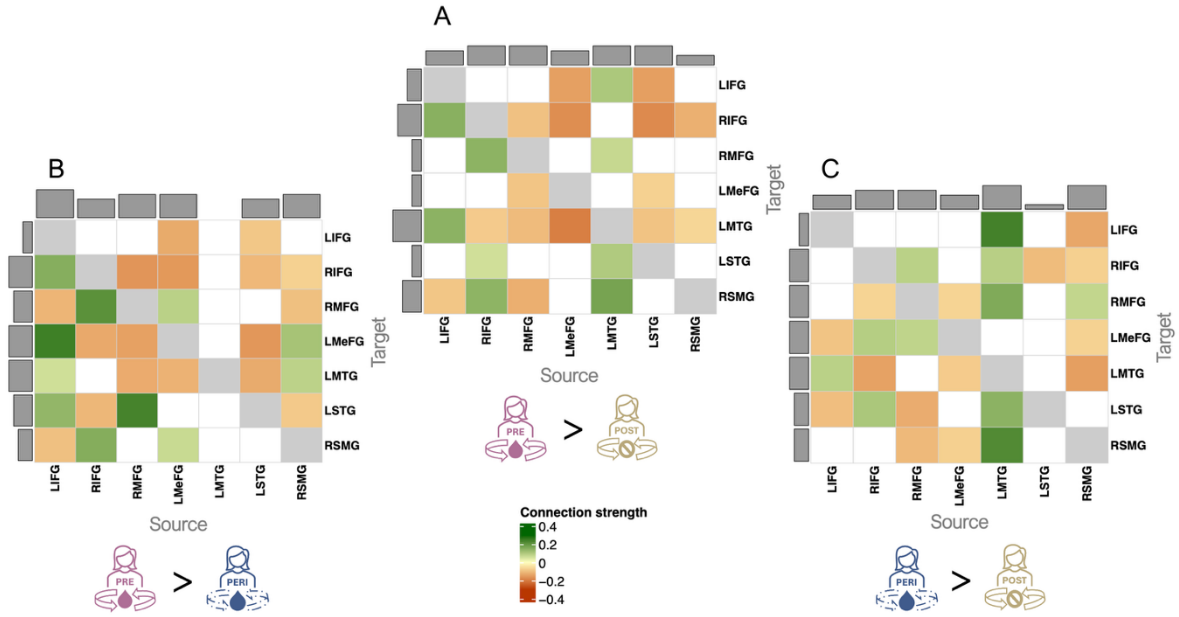


*Note*. All connections exceeding the posterior-probability criterion (PP > .95) are displayed, without applying a connectivity-strength threshold. Effects reflect pairwise comparisons from PEB models including age as a covariate (PRE > POST, PRE > PERI, PERI > POST). Abbreviations: LIFG, left inferior frontal gyrus; RIFG, right inferior frontal gyrus; RMFG, right middle frontal gyrus; LMeFG, left medial frontal gyrus; LMTG, left middle temporal gyrus; LSTG, left superior temporal gyrus; RSMG, right supramarginal gyrus.

**Supplementary Table S1.** Full set of connections with strong evidence (posterior probability > .95) for group differences in effective connectivity without applying a connectivity-strength threshold (age-corrected models).

| | Direction of connectivity | | | | |
|---|---|---|---|---|---|
| Group comparison | Source | | Target | Group comparison | Effect size in Hz |
| ***PRE > PERI*** | | | | | |
| *Inhibition* | | | | | |
| | RIFG | → | RMFG | + | .213 |
| | RIFG | → | LSTG | - | .110 |
| | RIFG | → | RSMG | + | .157 |
| | RMFG | → | LMeFG | - | -.145 |
| | RMFG | → | LMTG | - | -.128 |
| | RMFG | → | LSTG | + | .239 |
| | LMeFG | → | LIFG | - | -.131 |
| | LMeFG | → | RIFG | - | -.157 |
| | LMeFG | → | RMFG | + | .088 |
| | LMeFG | → | LMTG | - | -.118 |
| | LMeFG | → | RSMG | + | .070 |
| | LSTG | → | LIFG | - | -.085 |
| | LSTG | → | RIFG | - | -.109 |
| | LSTG | → | LMTG | - | -.130 |
| | RSMG | → | RIFG | - | -.070 |
| | RSMG | → | LMTG | + | .085 |
| *Excitation* | | | | | |
| | LIFG | → | RIFG | + | .156 |
| | LIFG | → | RMFG | - | -.114 |
| | LIFG | → | LMeFG | + | .248 |
| | LIFG | → | LMTG | + | .061 |
| | LIFG | → | LSTG | + | .139 |
| | LIFG | → | RSMG | - | -.096 |
| | RIFG | → | LMeFG | - | -.132 |
| | RMFG | → | RIFG | - | -.163 |
| | LSTG | → | LMeFG | - | -.160 |
| | RSMG | → | RMFG | - | -.097 |
| | RSMG | → | LMeFG | + | .114 |
| | RSMG | → | LSTG | - | -.081 |
| ***PRE > POST*** | | | | | |
| *Inhibition* | | | | | |
| | RIFG | → | LMTG | - | -.079 |
| | RIFG | → | LSTG | + | .059 |
| | RMFG | → | RIFG | - | -.095 |
| | RMFG | → | LMeFG | - | -.088 |
| | RMFG | → | LMTG | - | -.104 |
| | LMeFG | → | LIFG | - | -.146 |
| | LMeFG | → | RIFG | - | -.174 |
| | LMeFG | → | LMTG | - | -.198 |
| | LMTG | → | LIFG | + | .106 |

| | | | | | |
|---|---|---|---|---|---|
| | LMTG | → | RMFG | + | .073 |
| | LMTG | → | LSTG | + | .101 |
| | LSTG | → | LIFG | - | -.148 |
| | LSTG | → | RIFG | - | -.180 |
| | RSMG | → | RIFG | - | -.121 |
| *Excitation* | | | | | |
| | LIFG | → | RIFG | + | .152 |
| | LIFG | → | LMTG | + | .147 |
| | LIFG | → | RSMG | - | -.087 |
| | RIFG | → | RMFG | + | .146 |
| | RIFG | → | RSMG | + | .145 |
| | RMFG | → | RSMG | - | -.123 |
| | LMTG | → | RSMG | + | .177 |
| | LSTG | → | LMeFG | - | -.071 |
| | LSTG | → | LMTG | - | -.092 |
| | RSMG | → | LMTG | - | -.063 |
| ***PERI > POST*** | | | | | |
| *Inhibition* | | | | | |
| | LIFG | → | LMeFG | - | -.091 |
| | RIFG | → | RMFG | - | -.066 |
| | RIFG | → | LMeFG | + | .094 |
| | RIFG | → | LMTG | - | -.143 |
| | RIFG | → | LSTG | + | .108 |
| | RMFG | → | RIFG | + | .087 |
| | RMFG | → | LMeFG | + | .081 |
| | RMFG | → | LSTG | - | -.126 |
| | LMeFG | → | RMFG | - | -.066 |
| | LMeFG | → | LMTG | - | -.078 |
| | LMeFG | → | RSMG | - | -.072 |
| | LMTG | → | LIFG | + | .243 |
| | LMTG | → | RIFG | + | .092 |
| | LMTG | → | LSTG | + | .147 |
| | LSTG | → | RIFG | - | -.103 |
| | RSMG | → | LIFG | - | -.135 |
| | RSMG | → | LMTG | - | -.148 |
| *Excitation* | | | | | |
| | LIFG | → | LMTG | + | .088 |
| | LIFG | → | LSTG | - | -.100 |
| | RMFG | → | RSMG | - | -.108 |
| | LMTG | → | RMFG | + | .164 |
| | LMTG | → | RSMG | + | .225 |
| | RSMG | → | RIFG | - | -.072 |
| | RSMG | → | RMFG | + | .078 |
| | RSMG | → | LMeFG | - | -.069 |

*Note*. All parameters shown demonstrated a posterior probability > .95, no connectivity-strength threshold is applied. Positive values (+) indicate greater effective connectivity strength for Group A relative to Group B; negative values (-) indicate reduced connectivity strength for Group A relative to Group B. Reported effects reflect pairwise comparisons (PRE > PERI, PRE > POST, PERI > POST) from PEB models including age as a covariate. Results from models without age correction are provided in Supplementary Table S2. Abbreviations: LIFG, left inferior frontal gyrus; RIFG, right inferior frontal gyrus; RMFG, right middle frontal gyrus; LMeFG, left medial frontal gyrus; LMTG, left middle temporal gyrus; LSTG, left superior temporal gyrus; RSMG, right supramarginal gyrus.

**Supplementary Figure S4. Connections showing strong evidence (posterior probability > .95) for group differences in effective connectivity across menopausal transition in models without age as a covariate:** (A) PRE > POST, (B) PRE > PERI, and (C) PERI > POST.

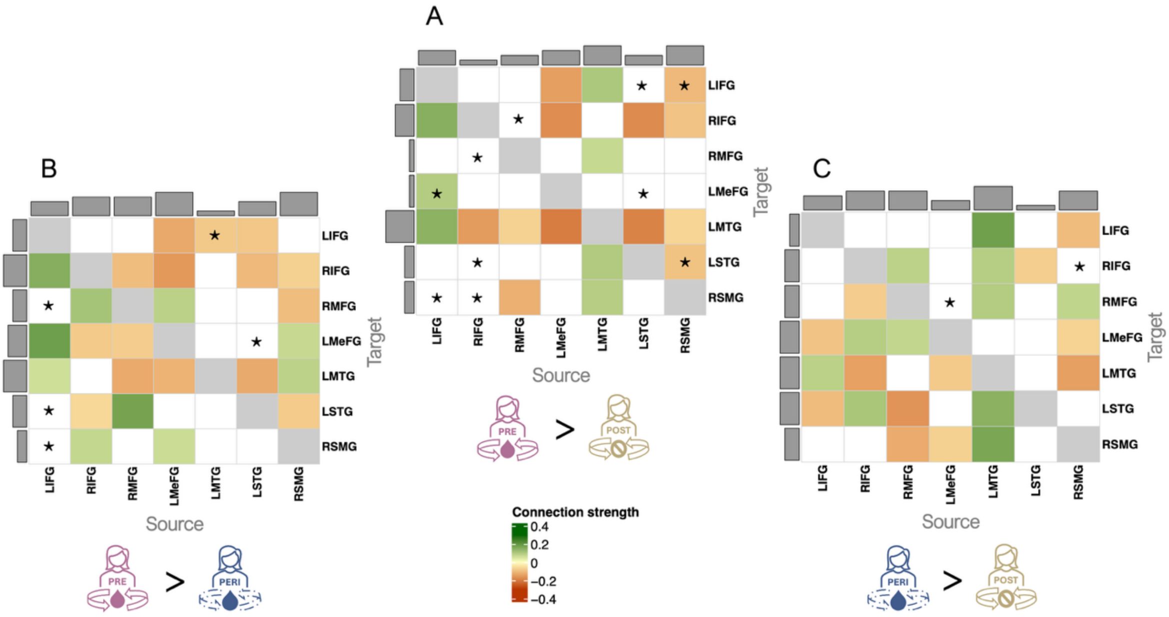


*Note*. All connections exceeding the posterior-probability criterion (PP > .95) are displayed. Effects reflect pairwise comparisons from PEB models estimated without age correction (PRE > POST, PRE > PERI, PERI > POST). Asterisks denote connections that appear or disappear when age is included as a covariate in the corresponding age-corrected models. Abbreviations: LIFG, left inferior frontal gyrus; RIFG, right inferior frontal gyrus; RMFG, right middle frontal gyrus; LMeFG, left medial frontal gyrus; LMTG, left middle temporal gyrus; LSTG, left superior temporal gyrus; RSMG, right supramarginal gyrus.

**Supplementary Table S2.** Full set of connections with strong evidence (posterior probability > .95) for group differences in effective connectivity in models without age as a covariate.

| | Direction of connectivity | | | | |
|---|---|---|---|---|---|
| Group comparison | Source | | Target | Group comparison | Effect size in Hz |
| ***PRE > PERI*** | | | | | |
| *Inhibition* | | | | | |
| | RIFG | → | RMFG | + | .114 |
| | RIFG | → | LSTG | - | -.060 |
| | RMFG | → | LMeFG | - | -.076 |
| | RMFG | → | LMTG | - | -.129 |
| | RMFG | → | LSTG | + | .177 |
| | LMeFG | → | LIFG | - | -.132 |
| | LMeFG | → | RIFG | - | -.157 |
| | LMeFG | → | RMFG | + | .087 |
| | LMeFG | → | LMTG | - | -.117 |
| | LMeFG | → | RSMG | + | .066 |
| | LSTG | → | LIFG | - | -.086 |
| | LSTG | → | RIFG | - | -.108 |
| | LSTG | → | LMTG | - | -.130 |
| | RSMG | → | RIFG | - | -.070 |
| | RSMG | → | LMTG | + | .085 |
| *Excitation* | | | | | |
| | LIFG | → | RIFG | + | .155 |
| | LIFG | → | LMeFG | + | .186 |
| | LIFG | → | LMTG | + | .061 |
| | RIFG | → | LMeFG | - | -.080 |
| | RIFG | → | RSMG | + | .076 |
| | RMFG | → | RIFG | - | -.101 |
| | LMTG | → | LIFG | - | -.081 |
| | RSMG | → | RMFG | - | -.100 |
| | RSMG | → | LMeFG | + | .072 |
| | RSMG | → | LSTG | - | -.078 |
| ***PRE > POST*** | | | | | |
| *Inhibition* | | | | | |
| | RIFG | → | LMTG | - | -.152 |
| | RMFG | → | LMTG | - | -.069 |
| | LMeFG | → | LIFG | - | -.147 |
| | LMeFG | → | RIFG | - | -.175 |
| | LMeFG | → | LMTG | - | -.200 |
| | LMTG | → | LIFG | + | .106 |
| | LMTG | → | RMFG | + | .073 |
| | LMTG | → | LSTG | + | .100 |
| | LMTG | → | RSMG | + | .095 |
| | LSTG | → | RIFG | - | -.179 |
| | LSTG | → | LMTG | - | -.190 |
| | RSMG | → | LIFG | - | -.108 |
| *Excitation* | | | | | |
| | LIFG | → | RIFG | + | .154 |

| | | | | | |
|---|---|---|---|---|---|
| | LIFG | → | LMeFG | + | .097 |
| | LIFG | → | LMTG | + | .148 |
| | RMFG | → | RSMG | - | -.121 |
| | RSMG | → | RIFG | - | -.092 |
| | RSMG | → | LMTG | - | -.064 |
| | RSMG | → | LSTG | - | -.092 |
| ***PERI > POST*** | | | | | |
| *Inhibition* | | | | | |
| | RIFG | → | RMFG | - | -.076 |
| | RIFG | → | LMeFG | + | .094 |
| | RIFG | → | LMTG | - | -.144 |
| | RIFG | → | LSTG | + | .110 |
| | RMFG | → | RIFG | + | .085 |
| | RMFG | → | LMeFG | + | .077 |
| | RMFG | → | LSTG | - | -.164 |
| | LMeFG | → | LMTG | - | -.079 |
| | LMeFG | → | RSMG | - | -.071 |
| | LMTG | → | LIFG | + | .183 |
| | LMTG | → | RIFG | + | .094 |
| | LMTG | → | LSTG | + | .150 |
| | LSTG | → | RIFG | - | -.075 |
| | RSMG | → | LIFG | - | -.102 |
| | RSMG | → | LMTG | - | -.147 |
| *Excitation* | | | | | |
| | LIFG | → | LMeFG | - | -.090 |
| | LIFG | → | LMTG | + | .088 |
| | LIFG | → | LSTG | - | -.103 |
| | RMFG | → | RSMG | - | -.128 |
| | LMTG | → | RMFG | + | .097 |
| | LMTG | → | RSMG | + | .170 |
| | RSMG | → | RMFG | + | .080 |
| | RSMG | → | LMeFG | - | -.067 |

*Note.* All parameters shown demonstrated a posterior probability > .95, no connectivity-strength threshold is applied. Positive values (+) indicate greater effective connectivity strength for Group A relative to Group B; negative values (-) indicate reduced connectivity strength for Group A relative to Group B. Reported effects reflect pairwise comparisons (PRE > PERI, PRE > POST, PERI > POST) from PEB models without age as a covariate. Abbreviations: LIFG, left inferior frontal gyrus; RIFG, right inferior frontal gyrus; RMFG, right middle frontal gyrus; LMeFG, left medial frontal gyrus; LMTG, left middle temporal gyrus; LSTG, left superior temporal gyrus; RSMG, right supramarginal gyrus.

**Supplementary Figure S5. Associations between effective connectivity and self-reported ability in emotion regulation (Difficulties in Emotion Regulation Scale-16, DERS-16):** (A) Connections showing strong evidence (posterior probability > .95) for associations with DERS-16 scores within each group, without applying a connectivity-strength threshold. (B) Leave-one-out cross-validation (LOOCV) results assessing the predictive validity of DERS-16 scores.

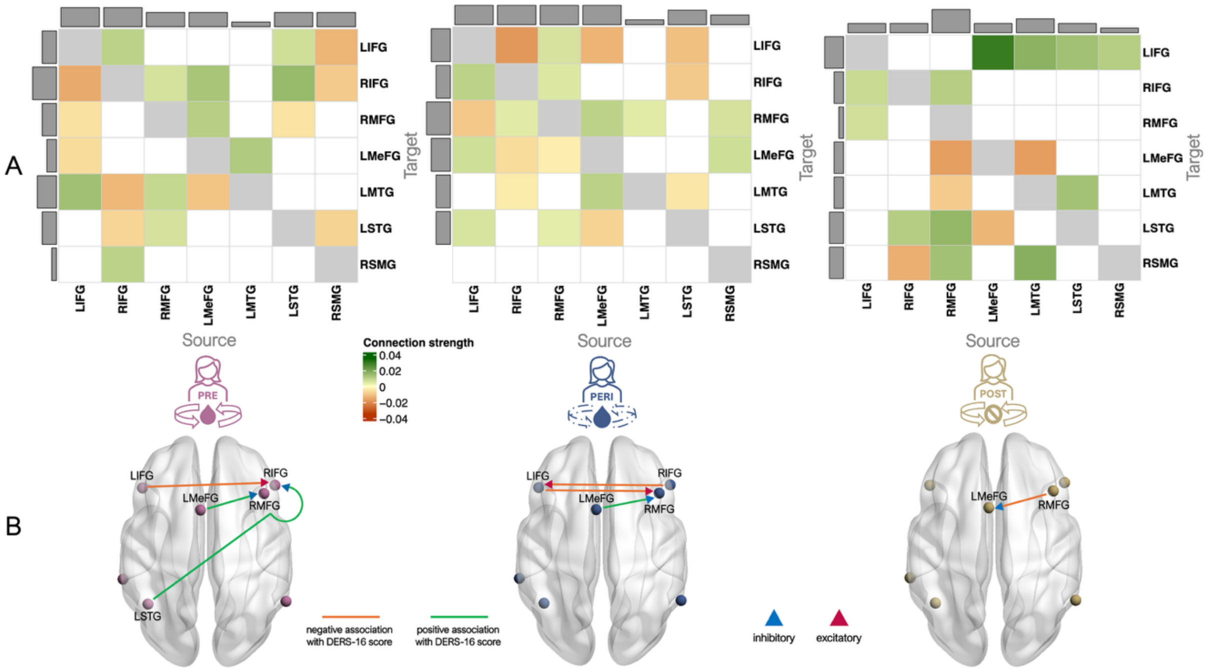


*Note*. Displayed connections in A exceed the posterior-probability criterion (PP > .95). Abbreviations: LIFG, left inferior frontal gyrus; RIFG, right inferior frontal gyrus; RMFG, right middle frontal gyrus; LMeFG, left medial frontal gyrus; LMTG, left middle temporal gyrus; LSTG, left superior temporal gyrus; RSMG, right supramarginal gyrus.

**Supplementary Table S3.** Full set of connections with strong evidence (posterior probability > .95) for associations between effective connectivity and self-reported emotion regulation ability (DERS-16) without applying a connectivity-strength threshold.

| | Direction of connectivity | | | | |
|---|---|---|---|---|---|
| **Group** | **Source** | | **Target** | **Relation with DERS-16** | **Effect size in Hz** |
| ***PRE*** | | | | | |
| *Inhibition* | | | | | |
| | RIFG | → | LMTG | - | -.015 |
| | RIFG | → | LSTG | - | -.008 |
| | RMFG | → | RIFG | + | .007 |
| | RMFG | → | LMTG | + | .010 |
| | RMFG | → | LSTG | + | .007 |
| | LMeFG | → | RIFG | + | .015 |
| | LMeFG | → | RMFG | + | .012 |
| | LMeFG | → | LMTG | - | -.012 |
| | LMTG | → | LMeFG | + | .014 |
| | LSTG | → | LIFG | + | .008 |
| | LSTG | → | RIFG | + | .018 |
| | RSMG | → | LIFG | - | -.015 |
| | RSMG | → | RIFG | - | -.009 |
| | RSMG | → | LSTG | - | -.011 |
| *Excitation* | | | | | |
| | LIFG | → | RIFG | - | -.017 |
| | LIFG | → | RMFG | - | -.006 |
| | LIFG | → | LMeFG | - | -.007 |
| | LIFG | → | LMTG | + | .016 |
| | RIFG | → | LIFG | + | .011 |
| | RIFG | → | RSMG | + | .011 |
| | LSTG | → | RMFG | - | -.006 |
| ***PERI*** | | | | | |
| *Inhibition* | | | | | |
| | LIFG | → | LMeFG | + | .008 |
| | RIFG | → | RMFG | + | .005 |
| | RIFG | → | LMTG | - | -.004 |
| | RMFG | → | LIFG | + | .007 |
| | RMFG | → | LMeFG | - | -.003 |
| | RMFG | → | LSTG | + | .005 |
| | LMeFG | → | LIFG | - | -.015 |
| | LMeFG | → | RMFG | + | .011 |
| | LMeFG | → | LMTG | + | .012 |
| | LMeFG | → | LSTG | - | -.009 |
| | LSTG | → | LIFG | - | -.013 |
| | LSTG | → | RIFG | - | -.012 |
| *Excitation* | | | | | |
| | LIFG | → | RIFG | + | .012 |
| | LIFG | → | RMFG | - | -.011 |
| | LIFG | → | LSTG | + | .007 |
| | RIFG | → | LIFG | - | -.021 |
| | RIFG | → | LMeFG | - | -.007 |

| | | | | | |
|---|---|---|---|---|---|
| | RMFG | → | RIFG | + | .006 |
| | LMTG | → | RMFG | + | .005 |
| | LSTG | → | LMTG | - | -.005 |
| | RSMG | → | RMFG | + | .007 |
| | RSMG | → | LMeFG | + | .008 |
| ***POST*** | | | | | |
| *Inhibition* | | | | | |
| | RIFG | → | LSTG | + | .013 |
| | RMFG | → | RIFG | + | .013 |
| | RMFG | → | LMeFG | - | -.020 |
| | LMeFG | → | LIFG | + | .034 |
| | LMeFG | → | LSTG | - | -.015 |
| | LMTG | → | LIFG | + | .020 |
| | LMTG | → | RSMG | + | .020 |
| | LSTG | → | LIFG | + | .016 |
| *Excitation* | | | | | |
| | LIFG | → | RIFG | + | .009 |
| | LIFG | → | RMFG | + | .008 |
| | RIFG | → | RSMG | - | -.016 |
| | RMFG | → | LMTG | - | -.010 |
| | RMFG | → | LSTG | + | .018 |
| | RMFG | → | RSMG | + | .016 |
| | LMTG | → | LMeFG | - | -.020 |
| | LSTG | → | LMTG | + | .016 |
| | RSMG | → | LIFG | + | .013 |

*Note*. All parameters shown demonstrated a posterior probability > .95, no connectivity-strength threshold is applied. Positive values (+) indicate a positive association between effective connectivity strength and DERS-16 scores, whereas negative values (-) indicate a negative association. Abbreviations: LIFG, left inferior frontal gyrus; RIFG, right inferior frontal gyrus; RMFG, right middle frontal gyrus; LMeFG, left medial frontal gyrus; LMTG, left middle temporal gyrus; LSTG, left superior temporal gyrus; RSMG, right supramarginal gyrus.
